# Development and Multinational Validation of a Novel Algorithmic Strategy for High Lp(a) Screening

**DOI:** 10.1101/2023.09.18.23295745

**Authors:** Arya Aminorroaya, Lovedeep S Dhingra, Evangelos K Oikonomou, Seyedmohammad Saadatagah, Phyllis Thangaraj, Sumukh Vasisht Shankar, Erica S Spatz, Rohan Khera

**Affiliations:** Section of Cardiovascular Medicine, Department of Internal Medicine, Yale School of Medicine, New Haven, CT, USA; Department of Medicine, Baylor College of Medicine, Houston, Texas, USA; Center for Outcomes Research and Evaluation (CORE), Yale New Haven Hospital, New Haven, CT, USA; Section of Health Informatics, Department of Biostatistics, Yale School of Public Health, New Haven, CT, USA

## Abstract

**Importance:** Elevated lipoprotein(a) [Lp(a)] is associated with atherosclerotic cardiovascular disease (ASCVD) and major adverse cardiovascular events (MACE). However, fewer than 0.5% of patients undergo Lp(a) testing, limiting the evaluation and use of novel targeted therapeutics currently under development.

**Objective:** We developed and validated a machine learning model to enable targeted screening for elevated Lp(a).

**Design:** Cross-sectional

**Setting:** 4 multinational population-based cohorts

**Participants:** We included 456,815 participants from the UK Biobank (UKB), the largest cohort with protocolized Lp(a) testing for model development. The model’s external validity was assessed in Atherosclerosis Risk in Communities (ARIC) (N=14,484), Coronary Artery Risk Development in Young Adults (CARDIA) (N=4,124), and Multi-Ethnic Study of Atherosclerosis (MESA) (N=4,672) cohorts.

**Exposures:** Demographics, medications, diagnoses, procedures, vitals, and laboratory measurements from UKB and linked electronic health records (EHR) were candidate input features to predict high Lp(a). We used the pooled cohort equations (PCE), an ASCVD risk marker, as a comparator to identify elevated Lp(a).

**Main Outcomes and Measures:** The main outcome was elevated Lp(a) (≥150 nmol/L), and the number-needed-to-test (NNT) to find one case with elevated Lp(a). We explored the association of the model’s prediction probabilities with all-cause and cardiovascular mortality, and MACE.

**Results:** The Algorithmic Risk Inspection for Screening Elevated Lp(a) (ARISE) used low-density lipoprotein cholesterol, statin use, triglycerides, high-density lipoprotein cholesterol, history of ASCVD, and anti-hypertensive medication use as input features. ARISE outperformed cardiovascular risk stratification through PCE for predicting elevated Lp(a) with a significantly lower NNT (4.0 versus 8.0 [with or without PCE], P<0.001). ARISE performed comparably across external validation cohorts and subgroups, reducing the NNT by up to 67.3%, depending on the probability threshold. Over a median follow-up of 4.2 years, a high ARISE probability was also associated with a greater hazard of all-cause death and MACE (age/sex-adjusted hazard ratio [aHR], 1.35, and 1.38, respectively, P<0.001), with a greater increase in cardiovascular mortality (aHR, 2.17, P<0.001).

**Conclusions and Relevance:** ARISE optimizes screening for elevated Lp(a) using commonly available clinical features. ARISE can be deployed in EHR and other settings to encourage greater Lp(a) testing and to improve identifying cases eligible for novel targeted therapeutics in trials.

**KEY POINTS:** *Question:* How can we optimize the identification of individuals with elevated lipoprotein(a) [Lp(a)] who may be eligible for novel targeted therapeutics?

*Findings:* Using 4 multinational population-based cohorts, we developed and validated a machine learning model, Algorithmic Risk Inspection for Screening Elevated Lp(a) (ARISE), to enable targeted screening for elevated Lp(a). In contrast to the pooled cohort equations that do not identify those with elevated Lp(a), ARISE reduces the “number-needed-to-test” to find one case with elevated Lp(a) by up to 67.3%.

*Meaning:* ARISE can be deployed in electronic health records and other settings to enable greater yield of Lp(a) testing, thereby improving the identification of individuals with elevated Lp(a).

## BACKGROUND

Lipoprotein(a) [Lp(a)] is associated with an increased risk of atherosclerotic cardiovascular disease (ASCVD) and major adverse cardiovascular events (MACE), even among individuals receiving optimal lipid-lowering therapy.^1–7^ Consequently, there have been broad recommendations for Lp(a) testing, with a single lifetime test sufficient due to stable serum levels from childhood.^8–12^ However, fewer than 0.5% of patients undergo Lp(a) testing based on contemporary real-world assessments.^13–15^ The underrecognition of those with an elevated Lp(a) is a pressing concern due to the rapidly evolving landscape for the Lp(a)-associated cardiovascular risk and its management.

Currently, individuals with elevated Lp(a) levels derive some benefit from counseling and rigorous cardiovascular risk factor modification but remain at elevated risk due to lack of targeted therapy.^11,12^ Novel Lp(a)-lowering agents, however, reduce serum Lp(a) by up to 97%,^16,17^ with several ongoing randomized controlled trials evaluating their safety and efficacy for cardiovascular risk reduction.^18,19^ These developments require the efficient identification of individuals with elevated Lp(a). Based on population prevalence, 1 in 8 individuals have elevated serum Lp(a), thereby posing a large burden of testing before identifying one case who may be eligible for enrollment in these studies and may benefit from treatment.^7,20,21^ Therefore, a testing prioritization strategy can rapidly target groups for trial enrollment by identifying those more likely to have elevated Lp(a) levels.

In this study, we developed and externally validated a novel machine learning model using structured clinical elements to optimize screening for elevated Lp(a), which can be deployed in clinical settings for case identification.

## METHODS

### Data Sources

The model was developed in the UK Biobank (UKB) (research application #71033) and externally validated in large US-based prospective cohort studies of Atherosclerosis Risk in Communities (ARIC), Coronary Artery Risk Development in Young Adults (CARDIA), and Multi-Ethnic Study of Atherosclerosis (MESA) (**Figure 1**).

**Figure 1.**
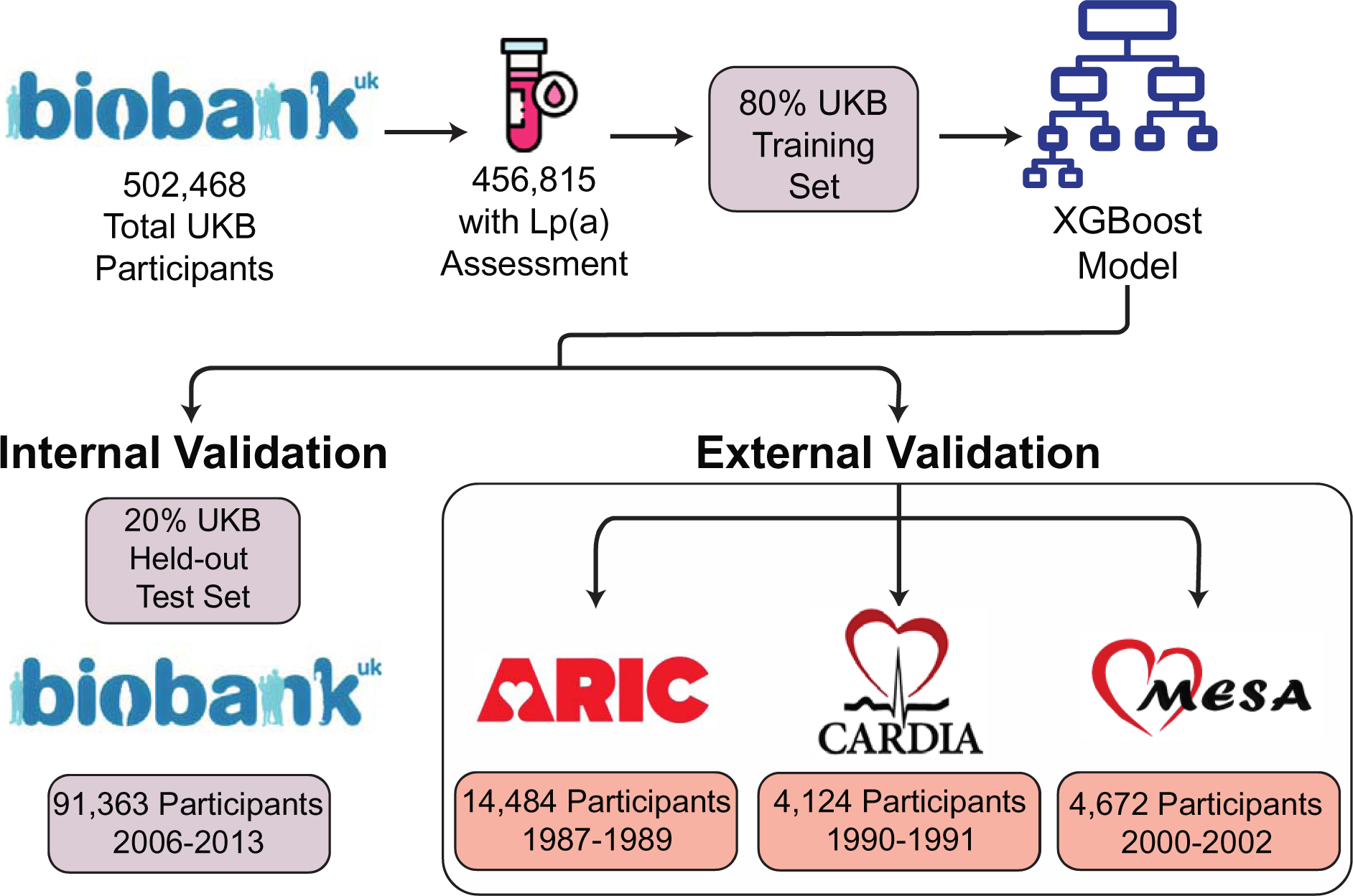
Study Design. Abbreviations: ARIC, Atherosclerosis Risk in Communities; CARDIA, Coronary Artery Risk Development in Young Adults; MESA, Multi-Ethnic Study of Atherosclerosis; UKB, UK Biobank.

We used the UKB comprising the largest cohort of individuals with protocolized Lp(a) assessment for model development.^22^ UKB is a prospective observational study of 502,468 people aged 40-69 years recruited in 2006-2010. In the baseline assessment, information across several domains was systematically collected, including medical history and laboratory values. All participants have linked their electronic health records (EHR), including hospital inpatient diagnoses and procedures. We included data from 456,815 (90.9%) participants with an Lp(a) assessment for model development (**eFigure 1**).

The ARIC study is a prospective observational study that enrolled 15,792 adults aged 45-64 years from four US communities using weighted sampling during 1987-1989.^23^ The ARIC study was designed to explore the etiology of atherosclerosis and the temporal evolution of cardiovascular risk factors and outcomes. In ARIC, 14,484 adults had Lp(a) measurement using their baseline blood sample.

In the CARDIA study, 5,116 adults aged 18-30 years were recruited from 4 US urban areas during 1985-1986 with prospective follow-up visits to examine the factors associated with the development of coronary artery disease in young adults.^24^ For validation, we included 4,124 participants who had undergone Lp(a) measurement during their third visit in 1990-1991.

MESA is a population-based study that enrolled 6,814 US adults aged 45-84 years from 6 centers during 2000-2002.^25^ The MESA study aimed to investigate the prevalence and evolution of subclinical ASCVD in a multi-ethnic cohort of Asian, Black, Chinese, and White individuals, where 4,672 underwent Lp(a) measurement at their baseline assessment.

### Study Outcome

The study outcome was an elevated serum Lp(a) defined as ≥150 nmol/L, representing the Lp(a) threshold for individuals enrolled in randomized clinical trials of Lp(a)-lowering agents.^19^ We reported the model’s number needed to test (NNT) to find one case with elevated Lp(a). Details for Lp(a) measurement across different cohorts are described in **eMethods**.

### Study Covariates

We defined a comprehensive set of potential covariates for model development, spanning demographics, vitals, diagnoses, procedures, medication history, and laboratory values drawn from the UKB study visits and linked data from the national UK EHR, including records predating UKB recruitment. We also calculated 10-year ASCVD risk for UKB participants using the pooled cohort equations (PCE), which is a readily available tool for ASCVD risk stratification, although not specifically designed for predicting high Lp(a), as a comparator.^26^ A detailed summary of the study covariates is presented in **eMethods**.

### Model Development

For model development, the UKB cohort was divided into train and held-out test sets (80:20 ratio), where the test set was not used in either model development or fine-tuning (**Figure 1**). All covariates were included as candidate predictors. Among these, all pairwise correlations were evaluated for all covariates to each other and to Lp(a). Among those covariates with a pairwise correlation of higher than 0.8, only one was retained to avoid collinearity, and the chosen covariate from the pair was the one with a higher correlation with serum Lp(a). Missing features were imputed using 5 nearest neighbors using the k-nearest neighbors algorithm.

Numerical features were scaled to achieve a zero mean and unit variance, while categorical features with more than two states were one-hot encoded. To develop the most efficient model, we employed both conventional machine learning models and deep learning methods, including logistic regression, extreme gradient boosting (XGBoost), and TabNet, a deep tabular data learning architecture.^27^ We trained multiple models using these architectures and all available features to achieve the best discrimination, assessed by the area under the receiver operating characteristic (AUROC). To select the final model, we prioritized low NNT and high AUROC while minimizing the number of features using cross-validation. We used SHapley Additive exPlanations (SHAP) values to determine the contribution of each feature to the model’s prediction probabilities for feature selection. The model’s hyperparameters were optimized for each model using five-fold cross-validation by grid search. For the final model, we reported the NNT to find one case with high Lp(a) in the held-out test set and external validation cohorts of ARIC, CARDIA, and MESA.

### Association with Clinical Outcomes

We evaluated whether the model identified the appropriate risk profile for inclusion in the studies, represented by the association of ARISE with outcomes. For this, we evaluated all-cause and cardiovascular death, and MACE (composite of acute coronary syndrome, ischemic stroke, and all-cause death) across the model’s prediction groups of high versus low probability of elevated Lp(a). Deaths were ascertained from linked death registries of England & Wales and Scotland from 2006 onwards (**eTable 1**).

We also evaluated the outcome benefit of Lp(a) assessment in those with a high probability of Lp(a). For this, we compared all-cause death and MACE rates between those who underwent Lp(a) testing and those who did not, across groups with high and low probability of elevated Lp(a) defined by the model. As noted in the data sources section, 9.1% of UKB participants did not undergo Lp(a) assessment. These were combined with the held-out test set as the group with Lp(a) assessment for these analyses. While the aliquots for Lp(a) assessment were drawn at the first or second study visit of the UKB during 2006-2013, Lp(a) assays were conducted during 2015-2017. Therefore, we used the Lp(a) assay date as the starting point for follow-up in the survival analyses, substituting missing dates with the median Lp(a) assay date.

### Statistical Analysis

Continuous and categorical variables were described as mean (standard deviation [SD]), and number (%), respectively. We evaluated the model’s performance using AUROC and area under the precision-recall curve (AUPRC) in the UKB and external validation cohorts. We also reported sensitivity, specificity, positive predictive value (PPV), negative predictive value (NPV), and F1 score across different probability thresholds based on optimizing specificity, the F1 score, and Youden’s index (maximizing sensitivity and specificity). The overall NNT, which represents the number of individuals tested for each person with elevated Lp(a), was calculated as the reciprocal of the prevalence of high Lp(a). For each model, the NNT was calculated as the reciprocal of the model’s PPV. We also computed the relative reduction of NNT using the model.

Finally, we calculated 95% confidence interval (CI) for AUROC and AUPRC using bootstrapping by resampling with replacement, while 95% CI for other performance measures was calculated using the standard error formula for proportion. To explore the association between the model’s prediction probability with mortality, we used age- and sex-adjusted Cox proportional hazard models. We used the model’s prediction groups and whether the patient received Lp(a) testing as independent variables to predict time-to-death. Statistical analyses were conducted using Python 3.11.2, and R version 4.2.0 with the statistical significance level set at 0.05.

## RESULTS

### Study Population

Of 502,468 UKB participants, 456,815 (90.9%) underwent Lp(a) assessment. The baseline characteristics of those with and without Lp(a) assessment were similar (**eFigure 1**, **eTable 2**). Participants in the development set had a mean age of 57.2 (8.1) years and 247,511 (54.2%) were women. Of those tested, 57,352 (12.6%) individuals had an elevated serum Lp(a). Compared with those with normal Lp(a), people with high Lp(a) were more likely to be women and Black and to have hypertension, ASCVD, family history of ASCVD, and premature ASCVD. High Lp(a) was associated with higher use of aspirin, statins, lipid-lowering therapies, and anti-hypertensive medications (**eTable 3**).

### Model Selection and Performance

We evaluated up to 4,575 features across three model architectures and selected an XGBoost model with the best performance in predicting elevated Lp(a) (**eTable 4**). This model, the Algorithmic Risk Inspection for Screening Elevated Lp(a) (ARISE), predicts elevated Lp(a) using six input features of ASCVD history, serum low-density lipoprotein cholesterol (LDL-C), serum high-density lipoprotein cholesterol (HDL-C), serum triglycerides, statin use, and anti-hypertensive medication use (available at https://www.cards-lab.org/arise). LDL-C was the most important feature in ARISE followed by statin use, triglycerides, HDL-C, ASCVD history, and anti-hypertensive medication use (**eFigure 2**).

**Figure 2.**
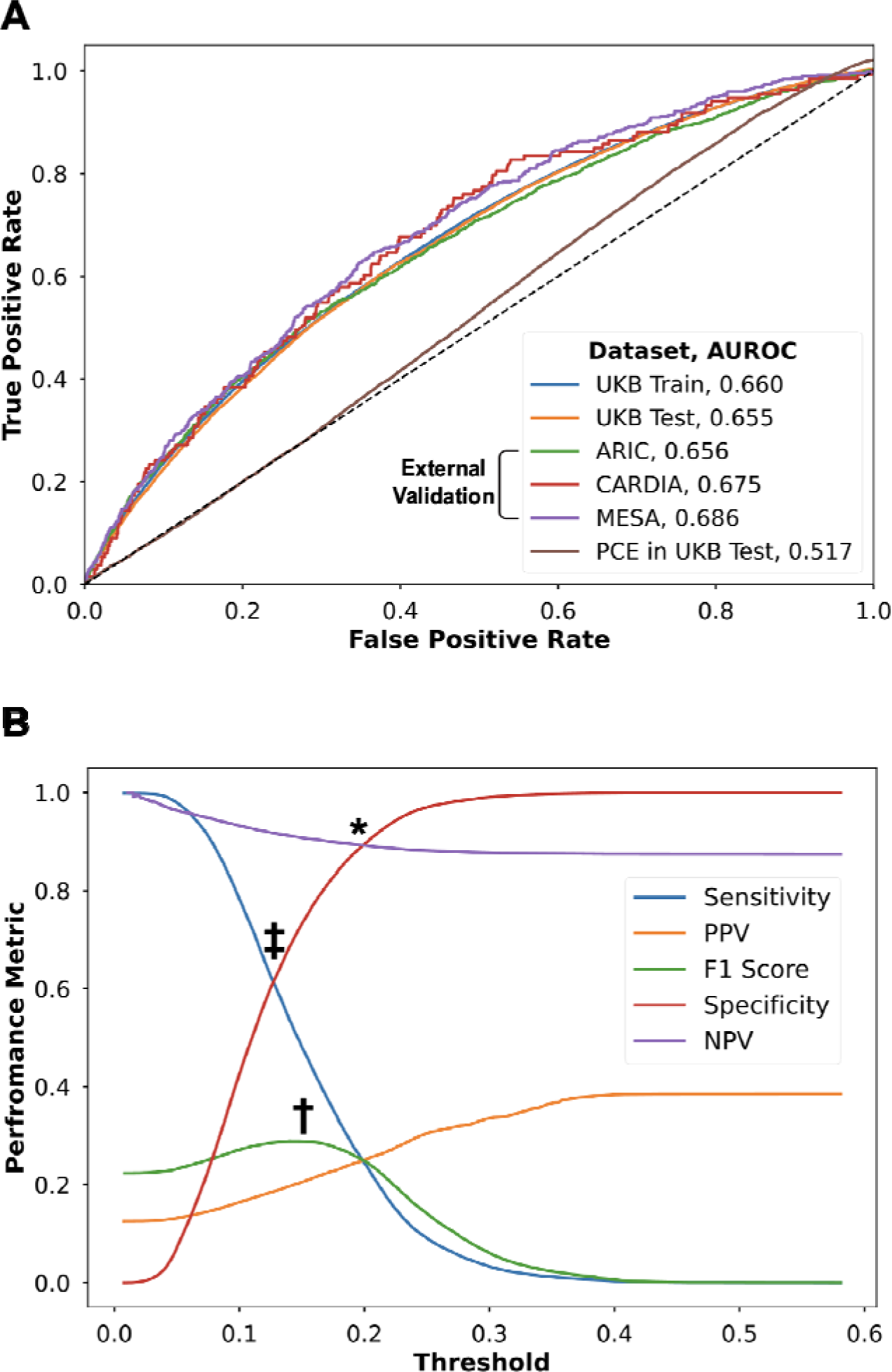
(A) Receiver Operating Characteristic Curves for the ARISE Across Study Cohorts and for the Pooled Cohort Equation in the UK Biobank Held-out Test Set; (B) ARISE’s Performance Measures Across Thresholds in the UK Biobank Held-out Test Set. *Optimized specificity at 90% †Optimized F1 score (maximizing sensitivity and PPV) ‡Optimized Youden’s index (maximizing sensitivity and specificity) Abbreviations: ARIC, Atherosclerosis Risk in Communities; AUROC, area under the receiver operating characteristic curve; CARDIA, Coronary Artery Risk Development in Young Adults; MESA, Multi-Ethnic Study of Atherosclerosis; NPV, negative predictive value; PCE, pooled cohort equation; PPV, positive predictive value; UKB, UK Biobank.

ARISE achieved an AUROC of 0.655 (95% CI, 0.650-0.661) and an AUPRC of 0.206 (95% CI, 0.200-0.212) in the held-out test set (**Figure 2A**, **Table 1**). The use of an ARISE-driven approach could reduce NNT by up to 67.3% from 8.0 without selective testing to 2.6, depending on the model’s probability threshold (**Figure 2B**). At a probability threshold for elevated Lp(a) of 0.203 for optimizing specificity at 90%, ARISE has a PPV of 0.243, corresponding to an NNT of 4.1. Alternatively, at the threshold optimizing Youden’s index, ARISE has a sensitivity of 0.621, a specificity of 0.605, and a PPV of 0.184 with an NNT of 5.4 (**Table 1**).

**Table 1.** Prevalence of High Lp(a), ARISE’s Performance, and ARISE’s Performance Measures Across Three Thresholds for Optimizing Specificity, F1 Score, and Youden’s Index in UK Biobank Held-out Test Set, ARIC, CARDIA, and MESA Cohorts.

| Characteristic* | UK Biobank Held-out Test Set | ARIC | CARDIA | MESA |
| --- | --- | --- | --- | --- |
| **High Lp(a) Prevalence** | 11,463 (12.5%) | 1661 (11.5%) | 133 (3.2%) | 495 (10.6%) |
| **AUROC (95% CI)** | 0.655 (0.650-0.661) | 0.656 (0.644-0.671) | 0.675 (0.627-0.717) | 0.686 (0.661-0.709) |
| **AUPRC (95% CI)** | 0.206 (0.200-0.212) | 0.200 (0.186-0.216) | 0.063 (0.047-0.093) | 0.200 (0.174-0.232) |
| **Optimal Specificity Threshold** |  |  |  |  |
| Sensitivity | 0.223 (0.220-0.226) | 0.204 (0.198-0.211) | 0.060 (0.053-0.067) | 0.059 (0.052-0.065) |
| Specificity | 0.900 (0.898-0.902) | 0.923 (0.919-0.928) | 0.972 (0.967-0.977) | 0.981 (0.977-0.985) |
| PPV | 0.243 (0.240-0.246) | 0.257 (0.250-0.264) | 0.067 (0.060-0.075) | 0.264 (0.251-0.276) |
| NPV | 0.890 (0.888-0.892) | 0.900 (0.895-0.904) | 0.969 (0.963-0.974) | 0.898 (0.889-0.907) |
| Overall NNT | 8.0 | 8.7 | 31.0 | 9.4 |
| Model NNT | 4.1 | 3.9 | 14.9 | 3.8 |
| NNT Relative Reduction | 48.3% | 55.3% | 52.0% | 59.8% |
| **Optimal F1 Score Threshold** |  |  |  |  |
| Sensitivity | 0.535 (0.532-0.538) | 0.506 (0.498-0.514) | 0.278 (0.265-0.292) | 0.277 (0.264-0.290) |
| Specificity | 0.684 (0.681-0.687) | 0.723 (0.716-0.730) | 0.868 (0.858-0.879) | 0.893 (0.884-0.902) |
| PPV | 0.196 (0.193-0.198) | 0.191 (0.185-0.198) | 0.066 (0.058-0.073) | 0.235 (0.222-0.247) |
| NPV | 0.911 (0.909-0.913) | 0.919 (0.914-0.923) | 0.973 (0.968-0.978) | 0.912 (0.904-0.921) |
| Overall NNT | 8.0 | 8.7 | 31.0 | 9.4 |
| Model NNT | 5.1 | 5.2 | 15.2 | 4.3 |
| NNT Relative Reduction | 35.9% | 40.1% | 50.9% | 54.8% |
| **Optimal Youden's Index Threshold** |  |  |  |  |
| Sensitivity | 0.621 (0.617-0.624) | 0.573 (0.565-0.581) | 0.361 (0.346-0.376) | 0.347 (0.334-0.361) |
| Specificity | 0.605 (0.602-0.609) | 0.651 (0.643-0.659) | 0.830 (0.819-0.842) | 0.843 (0.833-0.854) |
| PPV | 0.184 (0.182-0.187) | 0.175 (0.169-0.182) | 0.066 (0.059-0.074) | 0.208 (0.196-0.220) |
| NPV | 0.918 (0.916-0.919) | 0.922 (0.917-0.926) | 0.975 (0.970-0.980) | 0.916 (0.908-0.924) |
| Overall NNT | 8.0 | 8.7 | 31.0 | 9.4 |
| Model NNT | 5.4 | 5.7 | 15.1 | 4.8 |
| NNT Relative Reduction | 31.8% | 34.6% | 51.3% | 49.1% |

\*Data are presented as No. (%), No., percentage%, or point estimate (95% CI).
Abbreviations: ARIC, Atherosclerosis Risk in Communities; AUPRC, area under the precision-recall curve; AUROC, area under the receiver operating characteristics curve; CARDIA, Coronary Artery Risk Development in Young Adults; MESA, Multi-Ethnic Study of Atherosclerosis; NNT, number needed to test; NPV, negative predictive value; PPV, positive predictive value.

### Performance of ARISE versus Pooled Cohort Equation

Among UKB participants without previous ASCVD, 10-year ASCVD risk by PCE was 8.6% in those with high Lp(a) compared with 8.4% among those with normal Lp(a) (**eTable 3**). In this population without pre-existing ASCVD, PCE risk was not predictive of elevated Lp(a) levels, with an AUROC of 0.517 (95% CI, 0.511-0.523), which was significantly lower than ARISE (0.651, 95% CI, 0.646-0.656, P<0.001) (**Figure 2A**). There was no significant effect of PCE on NNT for Lp(a) testing and was 8.0 for both PCE-guided and non-selective testing.

### External Validation

The prevalence of high Lp(a) was 11.5% in ARIC, 3.2% in CARDIA, and 10.6% in MESA compared with 12.6% in the UKB (**eTable 5**). CARDIA participants were younger than participants of UKB, ARIC, and MESA. While 1.6% of UKB participants were Black, a large proportion of the external validation cohorts were Black (25.0-48.2%). Hispanic individuals comprised 22.7% of MESA participants. Among UKB participants, 4.9% had ASCVD and 16.7% were using statin compared with 16.0% and 0.6% in ARIC, and 8.2% and 0.3% in CARDIA, respectively. No one had a history of ASCVD, or statin use in the MESA cohort (**eTables 6-8**). ARISE performed comparably in the external validation cohorts with an AUROC of 0.653 in ARIC, 0.667 in CARDIA, and 0.685 in MESA (**Figure 2A**, **Table 1**).

Across study cohorts, the PPV without ARISE, which represents the prevalence of high Lp(a), ranged from 0.032 in CARDIA to 0.125 in the UKB test set, corresponding to an NNT ranging from 31.0 to 8.0, respectively (**Table 1**). At the threshold for optimizing specificity at 90%, ARISE reduces NNT by 48.3% in the UKB test set, 52.0% in CARDIA, 55.3% in ARIC, and 59.8% in MESA compared with non-selective testing. This was consistent at the threshold for optimized Youden’s index, with an NNT reduction by ARISE varied from 31.8% in the UKB test set to 51.3% in CARDIA.

### ARISE’s Performance in Participant Subgroups

ARISE performed comparably across subgroups of demographics, medical history, and medication history in the held-out test set from UKB and the external validation cohorts (**Figure 3**, **eFigures 3-5**, **eTables 9-12**). Across study cohorts, ARISE’s AUROC ranged from 0.652-0.691 in women compared with 0.650-0.695 in men, and 0.643-0.656 in Black individuals compared with 0.643-0.784 in White individuals. ARISE’s AUROC was 0.689 (95% CI, 0.631-0.744) among Hispanic individuals in MESA.

**Figure 3.**
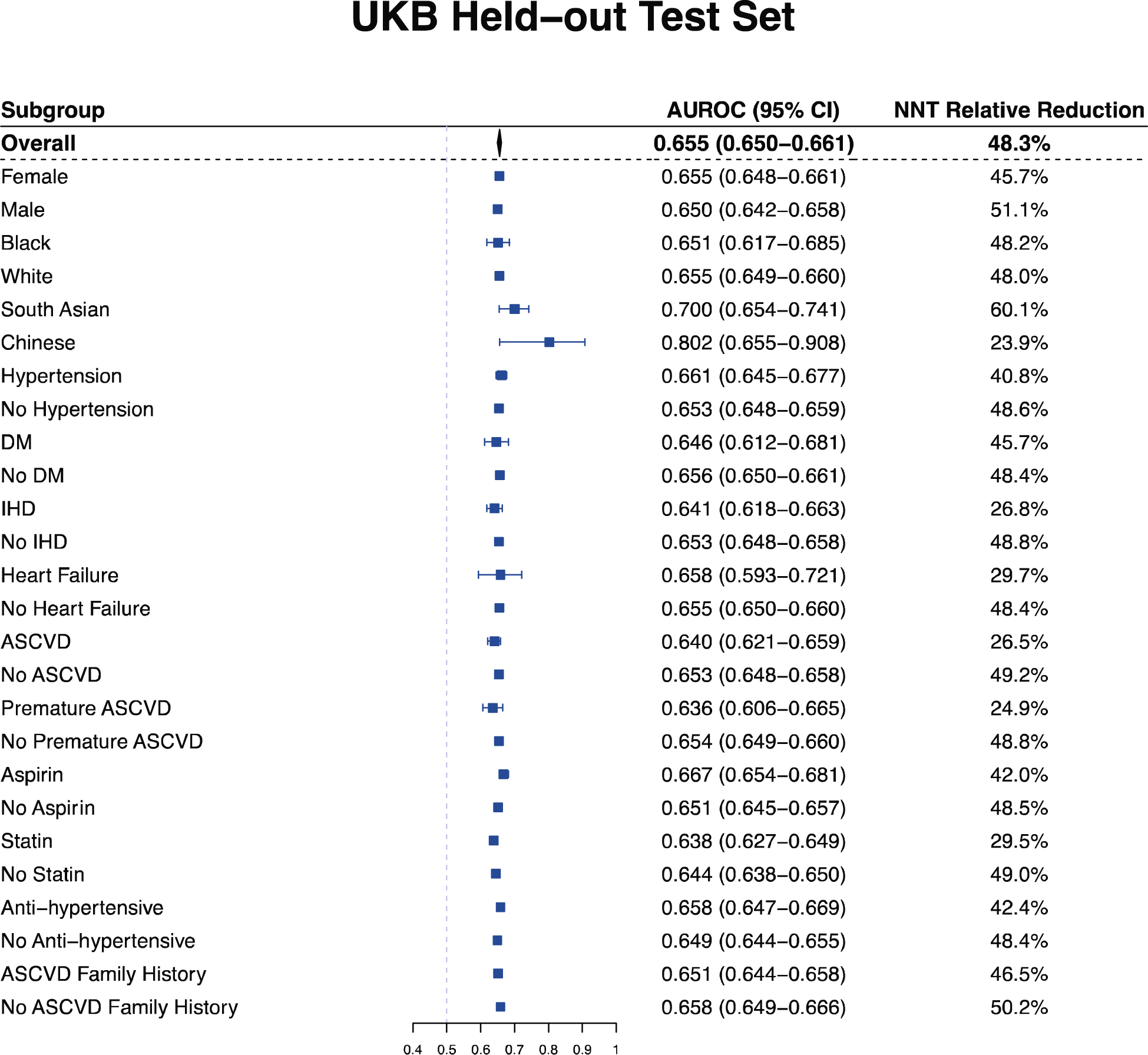
ARISE’s Performance and Number Needed to Test Relative Reduction Across Demographic and Clinical Subgroups in the UKB Held-out Test Set. Abbreviations: ASCVD, atherosclerotic cardiovascular disease; AUROC, area under the receiver operating characteristic curve; CI, confidence interval; DM, diabetes mellitus; IHD, ischemic heart disease; NNT, number needed to test; UKB, UK Biobank.

At the threshold for optimizing specificity, ARISE-optimized Lp(a) testing was associated with a 49.2%-57.5% reduction in NNT among people without ASCVD compared with 26.5%-42.3% among those with ASCVD in the UKB test set and ARIC. Of note, ARISE-optimization was associated with a larger NNT reduction for individuals without statin use and ASCVD family history compared with those with statin use and ASCVD family history, respectively, across cohorts (**eTables 9-12**).

### Outcome Analyses

Among 137,016 UKB participants, including 91,363 from the UKB test set with Lp(a) measurements and 45,653 UKB participants without Lp(a) measurements, 4,406 (3.3%) deaths, including 568 (0.4%) cardiovascular deaths, and 6,661 (5.1%) MACE occurred over a median follow-up of 4.2 years following the Lp(a) assay date (**eTable 13**). At the threshold for optimizing specificity, individuals with a high ARISE score were more likely to die and have a MACE compared with those with a low ARISE score (age/sex-adjusted hazard ratio [aHR], 1.35, 95% CI, 1.26-1.46, and aHR, 1.38, 95% CI, 1.30-1.46, respectively), with more pronounced increase in cardiovascular mortality (aHR, 2.17, 95% CI, 1.80-2.62) compared with non-cardiovascular mortality (aHR, 1.25, 95% CI, 1.15-1.36).

In the evaluation of outcome associations of Lp(a) assessment across strata of ARISE, participants from the UKB test set with Lp(a) measurements and UKB participants without Lp(a) measurements had similar baseline characteristics and included 17,438 (12.7%) participants with a high ARISE score (**eTable 13**). Among those with a high ARISE score, undergoing Lp(a) measurement was associated with a lower hazard of all-cause death and MACE compared with no Lp(a) testing (aHR, 0.82, 95% CI, 0.72-0.94, and aHR, 0.88, 95% CI, 0.79-0.99, respectively). However, the hazard of death and MACE was comparable between those with and without Lp(a) assessment among individuals with a low ARISE score (aHR, 0.95, 95% CI, 0.89-1.02, and aHR, 0.99, 95% CI, 0.93-1.05, respectively, **Figure 4**).

**Figure 4.**
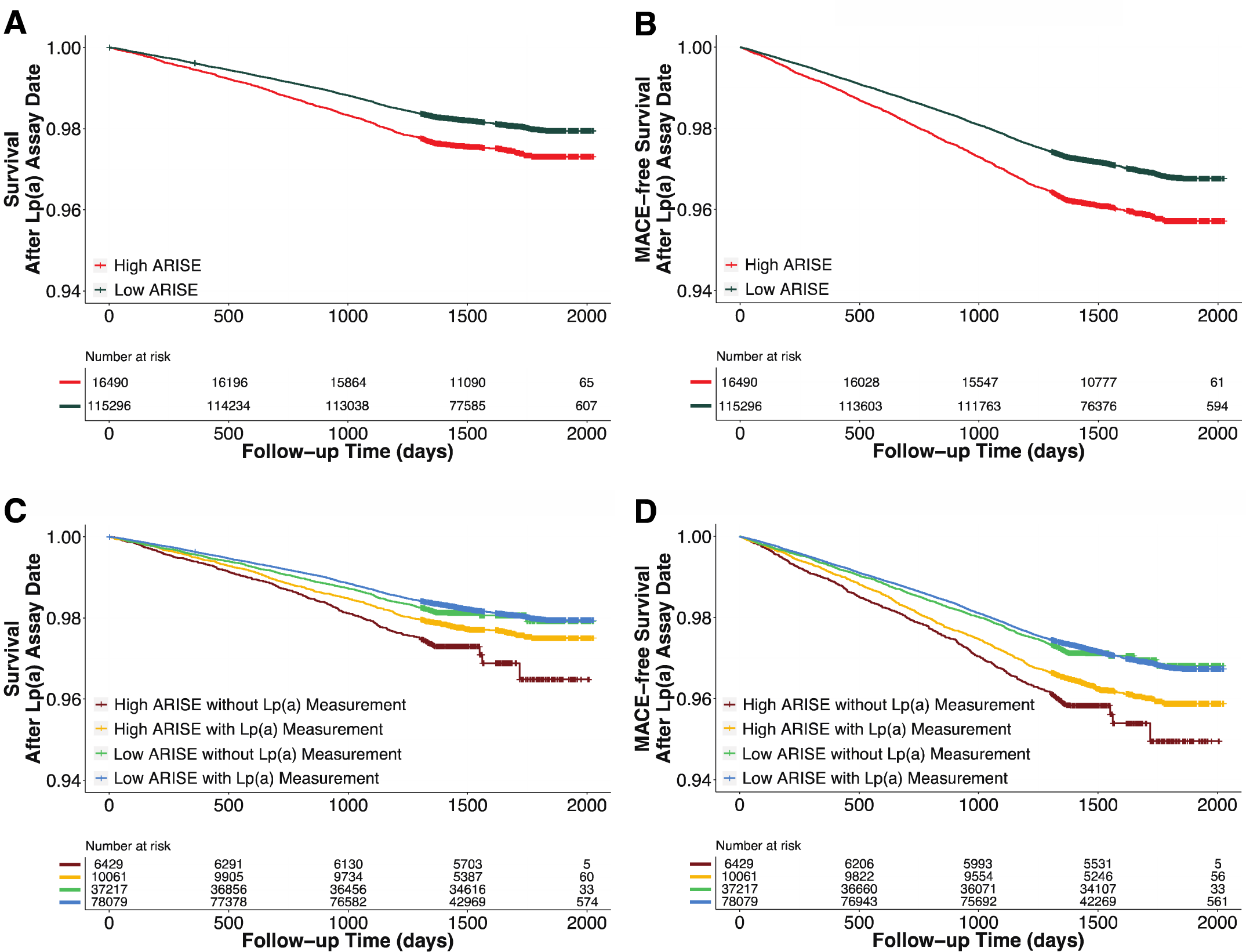
Age/Sex-adjusted (A) Survival and (B) MACE-free Survival Curves Across High and Low ARISE Scores; Age/Sex-adjusted (C) Survival and (D) MACE-free Survival Curves Across High and Low ARISE Scores Based on Lp(a) Assessment. Abbreviations: ARISE, Algorithmic Risk Inspection for Screening Elevated Lp(a); MACE, major adverse cardiovascular events.

## DISCUSSION

In the UKB, the largest cohort with protocolized Lp(a) assessment, we developed ARISE, a machine learning model that utilizes commonly available structured clinical elements available in EHRs to improve the yield of Lp(a) testing. ARISE significantly outperformed non-selective cardiovascular risk assessment (PCE) for predicting high Lp(a) and generalized well to the external validation cohorts of ARIC, CARDIA, and MESA. Across cohorts, the use of ARISE has the potential to increase the yield of Lp(a) testing by over 2-fold, with a more pronounced improvement among those without ASCVD compared with those with ASCVD.

The study builds upon the current literature by highlighting an algorithmic approach essential for enhancing the role of Lp(a) both as a prognostic marker and as a potential treatment target for elevated cardiovascular risk, while there are no alternative strategies for optimizing the identification of cases with elevated LP(a). Presently, the American Heart Association/American College of Cardiology (AHA/ACC) guidelines on the management of blood cholesterol recommend Lp(a) assessment as an ASCVD risk enhancer in select individuals with a family history of premature ASCVD.^28,29^ This approach does not cover broader population cardiovascular risk management, as nearly 10-15% live with an increased cardiovascular risk associated with high Lp(a).^7,20,21^ Given that ARISE reduces NNT to find those with high Lp(a), and one lifetime Lp(a) assessment is adequate, large-scale implementation of the model feasibly optimizes identifying patients with elevated Lp(a). Consequently, these patients can undergo rigorous cardiovascular risk management and initiation or intensification of existing therapies for elevated Lp(a), with the potential role of proprotein convertase subtilisin/kexin type 9 inhibitors in MACE reduction via lowering Lp(a).^30–34^

Clinical trials are underway to assess targeted therapies for lowering Lp(a).^18,19,35–38^ This underscores the importance of detecting people with high Lp(a), as therapies for elevated Lp(a) are currently available and targeted therapies are emerging. Since additional phase 3 trials are required to evaluate the safety and efficacy of the emerging therapeutics in primary and secondary prevention settings, ARISE-optimized Lp(a) testing holds promise to enable more efficient enrollment of these trials. Given that ARISE uses routinely collected EHR structured fields, its deployment within health systems for case finding can substantially increase the feasibility of conducting clinical trials for novel therapies targeting Lp(a).

We posit that systematic, algorithmic testing can be optimized to identify those at high cardiovascular risk, especially among those without pre-existing ASCVD, a group not explicitly highlighted in the current guidelines.^28,29^ Evidence from the UKB also suggests that individuals identified as high-risk for elevated Lp(a) using ARISE had better outcomes if they had a known Lp(a) level, as opposed to those without testing. However, this association was not observed in individuals with a lower ARISE score. This further reinforces the likely clinical utility of risk-optimized testing. Another key observation is that ARISE outperforms PCE, which may suggest that traditionally high-risk individuals, as commonly defined by PCE in clinical practice, would not be more likely to have elevated Lp(a). Therefore, a dedicated tool is essential to accomplish efficient testing for Lp(a)-driven risk. Finally, the consistent performance of ARISE across key demographic subgroups, despite known racial and ethnic variations in Lp(a), underscores that ARISE adoption holds potential for equitable improvement in patient outcomes.^7,39^

The study findings should be interpreted in the light of its limitations. First, ARISE does not have good discrimination or degree of predictions typical of high-performing predictive models. This reflects the largely independent and likely genetically determined risk from Lp(a) levels.^7^ Nevertheless, it outperforms standard risk stratification using PCE, and represents the best achievable performance using the wide range of structured clinical data in a high-quality cohort and leveraging state-of-the-art methods. Furthermore, ARISE can reduce the NNT by up to two-thirds and increase the yield of Lp(a) testing, a key advance without an existing alternative. Second, Lp(a) testing was not performed for some UKB participants. While this may be a concern for bias in model development, fewer than 10% of the UKB participants had no Lp(a) measurement, and those with and without Lp(a) measurements had similar baseline characteristics. Additionally, ARISE performed comparably across external cohorts. Third, ARISE was externally validated in ARIC, CARDIA, and MESA, where the prevalence of statin use was less than 1%. Nevertheless, the model performance was comparable across those with and without statin use in the UKB.

## CONCLUSIONS

We developed ARISE, a model for increasing the yield of Lp(a) testing using routinely available clinical features, which can be operationalized in the EHR. The use of an algorithmic approach to increase detection efficiency can accelerate discoveries in the therapeutic management of Lp(a)-related risk and improve cardiovascular outcomes of patients in the future.

## Supporting information

Supplemental Content

## Data Availability

The analyzed de-identified data are available for the UK Biobank cohort from the UK Biobank's Access Management System and for ARIC, CARDIA, and MESA cohorts from the NHLBI's Biologic Specimen and Data Repository Information Coordinating Center (BioLINCC). The additional supporting information (statistical/analytic code) is available upon request to the corresponding author.

https://ams.ukbiobank.ac.uk/ams/

https://biolincc.nhlbi.nih.gov/studies/

## Author Contributions

Drs Aminorroaya and Khera had full access to all of the data in the study and take responsibility for the integrity of the data and the accuracy of the data analysis. All authors approved the final version for submission.

*Study concept and design*: Aminorroaya, Arya; Khera, Rohan.

*Acquisition, analysis, or interpretation of data*: Aminorroaya, Arya; Dhingra, Lovedeep S; Oikonomou, Evangelos K; Saadatagah, Seyedmohammad; Thangaraj, Phyllis; Vasisht Shankar, Sumukh; Spatz, Erica; Khera, Rohan.

*Drafting of the manuscript*: Aminorroaya, Arya; Dhingra, Lovedeep S.

*Critical revision of the manuscript for important intellectual content*: Evangelos K; Saadatagah, Seyedmohammad; Thangaraj, Phyllis; Vasisht Shankar, Sumukh; Spatz, Erica; Khera, Rohan.

*Statistical analysis*: Aminorroaya, Arya.

*Obtained funding*: Khera, Rohan.

*Administrative, technical, or material support*: Aminorroaya, Arya; Dhingra, Lovedeep S; Vasisht Shankar, Sumukh; Khera, Rohan.

*Study supervision*: Khera, Rohan.

## Disclaimer

The views expressed in this article are those of the authors and not necessarily any funders.

