## Supplemental Content for "Development and Multinational Validation of a Novel Algorithmic Strategy for High Lp(a) Screening"

### **eMethods**

**Lp(a) Measurement Methodology**

In the UK Biobank (UKB), lipoprotein(a) [Lp(a)] was systematically measured using an immuno-turbidimetric assay by Randox Bioscience on a Beckman Coulter AU5800 platform and reported as nmol/L. While this assay may not be completely isoform insensitive, it utilizes the Denka Seiken method, which has demonstrated strong concordance with reference material from the International Federation of Clinical Chemistry and Laboratory Medicine.^1^

In Atherosclerosis Risk in Communities (ARIC), Lp(a) was measured using a double-antibody enzyme-linked immunosorbent assay (ELISA), which measures the Lp(a) protein moiety mass representing one-third of the total Lp(a) mass. Therefore, the reported values were adjusted by a correction factor of 3 to obtain the Lp(a) mass concentration, followed by the conversion factor of 2.15 for converting mg/dL to nmol/L to reach serum Lp(a) in nmol/L.^2–4^ In Coronary Artery Risk Development in Young Adults (CARDIA), Lp(a) mass concentration was determined using a double monoclonal antibody-based ELISA, and the reported values were converted to nmol/L by applying the conversion factor of 2.15.^4,5^ In Multi-Ethnic Study of Atherosclerosis (MESA), Lp(a) was assessed through a latex-enhanced turbidimetric immunoassay (Denka Seiken, Tokyo, Japan), and the results were reported in nmol/L.^1,6^

**Study Covariates**

Age, sex, ethnicity, smoking, body mass index, systolic and diastolic blood pressure, heart rate, family history, medication history, and laboratory values were collected during the UKB study visits. Family history of atherosclerotic cardiovascular disease (ASCVD) was defined as a history of stroke or heart disease in participants’ father, mother, or siblings. Medications were defined by participants’ self-report and validation by the UKB staff, and included aspirin, statin, lipid-lowering therapies, anti-hypertensive medications, insulin, hormone replacement therapy, and contraceptive pills (**eTable 1**).^7^ Candidate laboratory testing included complete blood count, lipid profile, blood sugar, hemoglobin A1C, liver enzymes and bilirubin, C-reactive protein, calcium, phosphate, total protein, albumin, and vitamin D, representing labs routinely collected in the clinical setting. The detailed methods for sample handling and conducting the measurements in the UKB have been previously described.^8,9^

We obtained diagnoses and procedures from UK EHR data, covering historical records predating UKB enrollment, including Hospital Episode Statistics for England (1997 onwards), Scottish Morbidity Record (1981 onwards), and Patient Episode Database for Wales (1998 onwards). Hypertension, diabetes mellitus, chronic kidney disease, ischemic heart disease (IHD), heart failure, ischemic stroke, and peripheral arterial disease (PAD) were identified by relevant International Classification of Diseases versions-9 and 10, ICD-9 and ICD-10 codes. Furthermore, the number of hospitalization episodes due to acute IHD, heart failure, ischemic stroke, and PAD was assessed for each participant (**eTable 1**). The history and number of procedures of coronary artery bypass grafting (CABG), percutaneous coronary intervention (PCI), carotid revascularization, and PAD revascularization were evaluated using the Office of Population Censuses and Surveys Classification of Interventions and Procedures, versions 3 and 4, OPCS-3 and OPCS-4 codes (**eTable 1**). In addition to the abovementioned conditions and procedures, each unique ICD and OPCS code was considered as a potential model feature for predicting high Lp(a).

Given the differences in serum Lp(a) based on the ASCVD history, we used ASCVD history, premature ASCVD history, and age at the first ASCVD event as potential input features for predicting high Lp(a).^10^ ASCVD was defined as a history of IHD, ischemic stroke, PAD, CABG, PCI, carotid revascularization, or PAD revascularization. Premature ASCVD was defined as the first episode of ASCVD in men <55 and women <65 years of age.

### **eFigure 1. Lp(a) Assessment among the UK Biobank Participants**

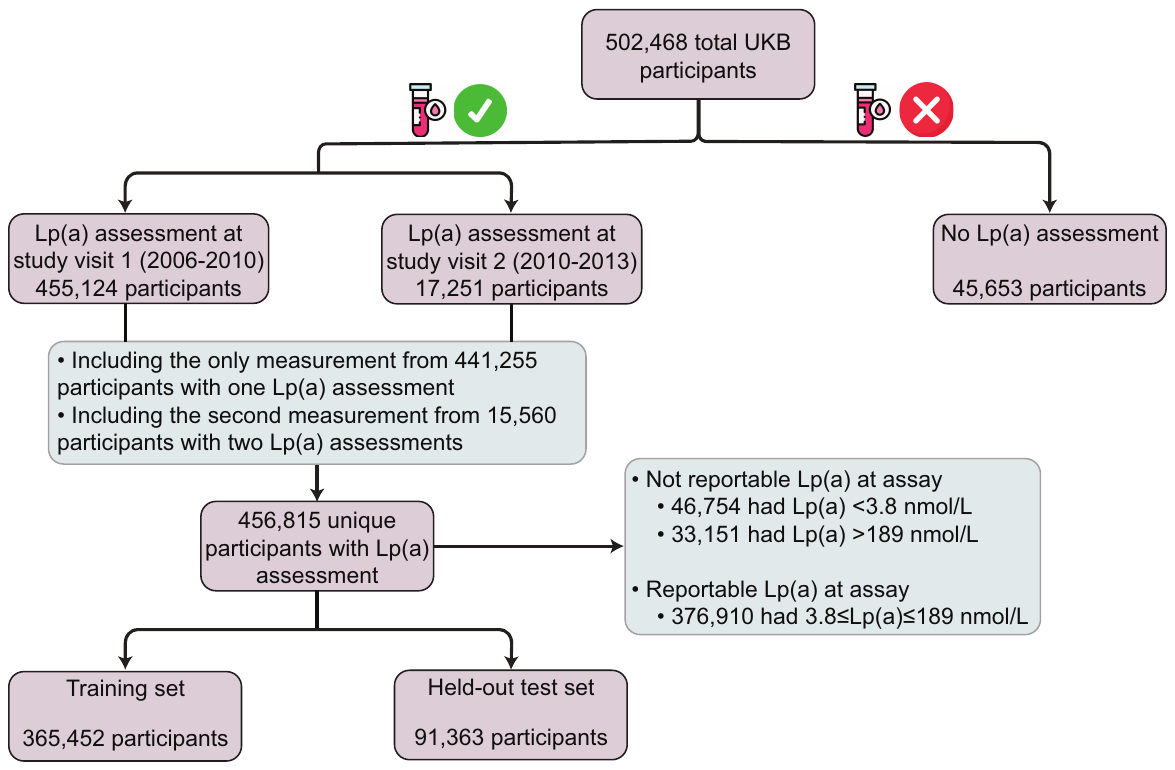

Abbreviations: UKB, UK Biobank.

### **eFigure 2. SHAP Values of ARISE’s Features Across UK Biobank Held-out Test Set, ARIC, CARDIA, and MESA Cohorts**

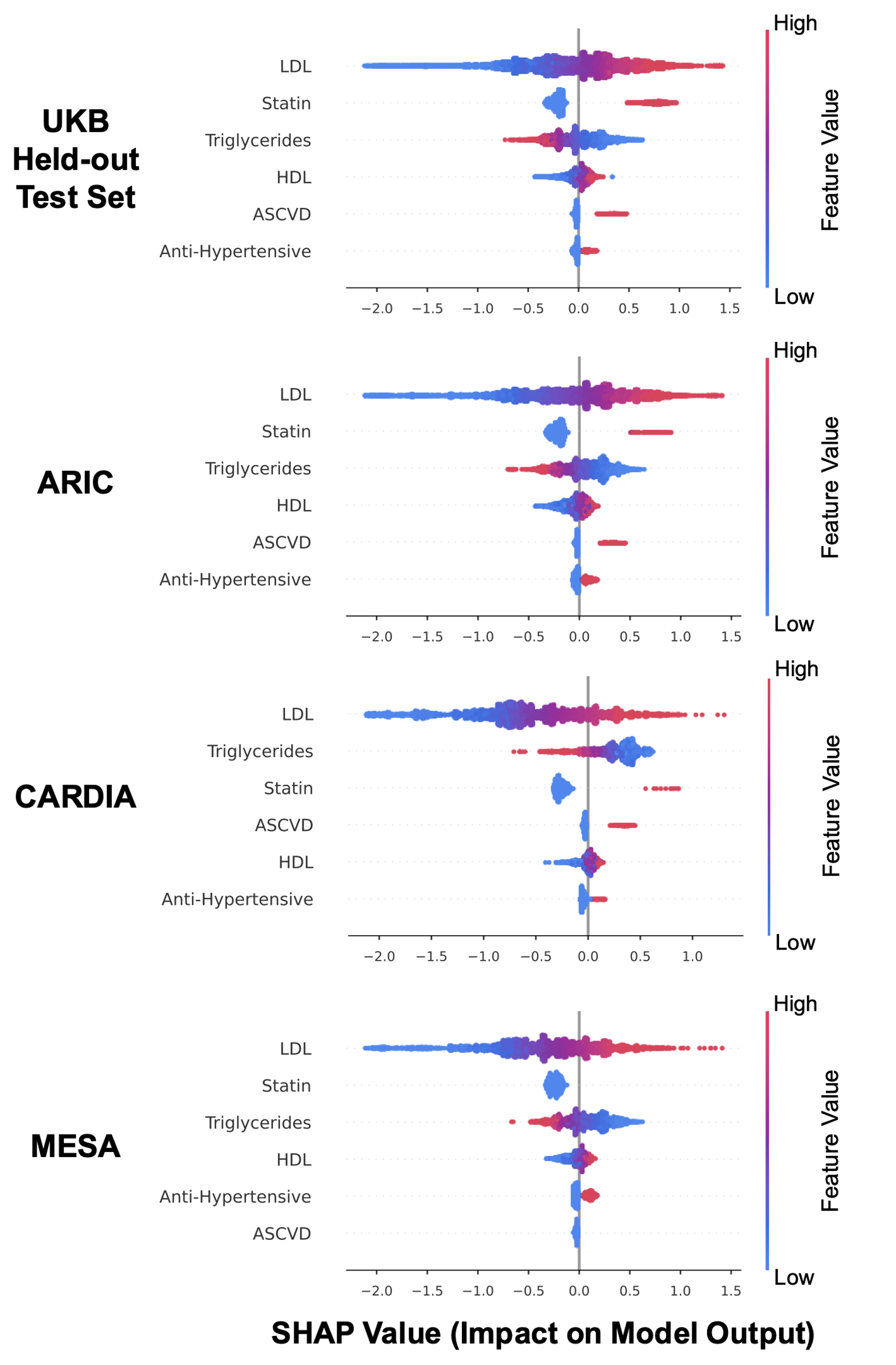

Abbreviations: ARIC, Atherosclerosis Risk in Communities; ASCVD, atherosclerotic cardiovascular disease; CARDIA, Coronary Artery Risk Development in Young Adults; HDL, high-density lipoprotein cholesterol; LDL, low-density lipoprotein cholesterol; MESA, Multi-Ethnic Study of Atherosclerosis; SHAP, SHapley Additive exPlanations; UKB, UK Biobank.

### **eFigure 3. ARISE’s Performance and Number Needed to Test Relative Reduction Across Demographic and Clinical Subgroups in the ARIC Cohort**

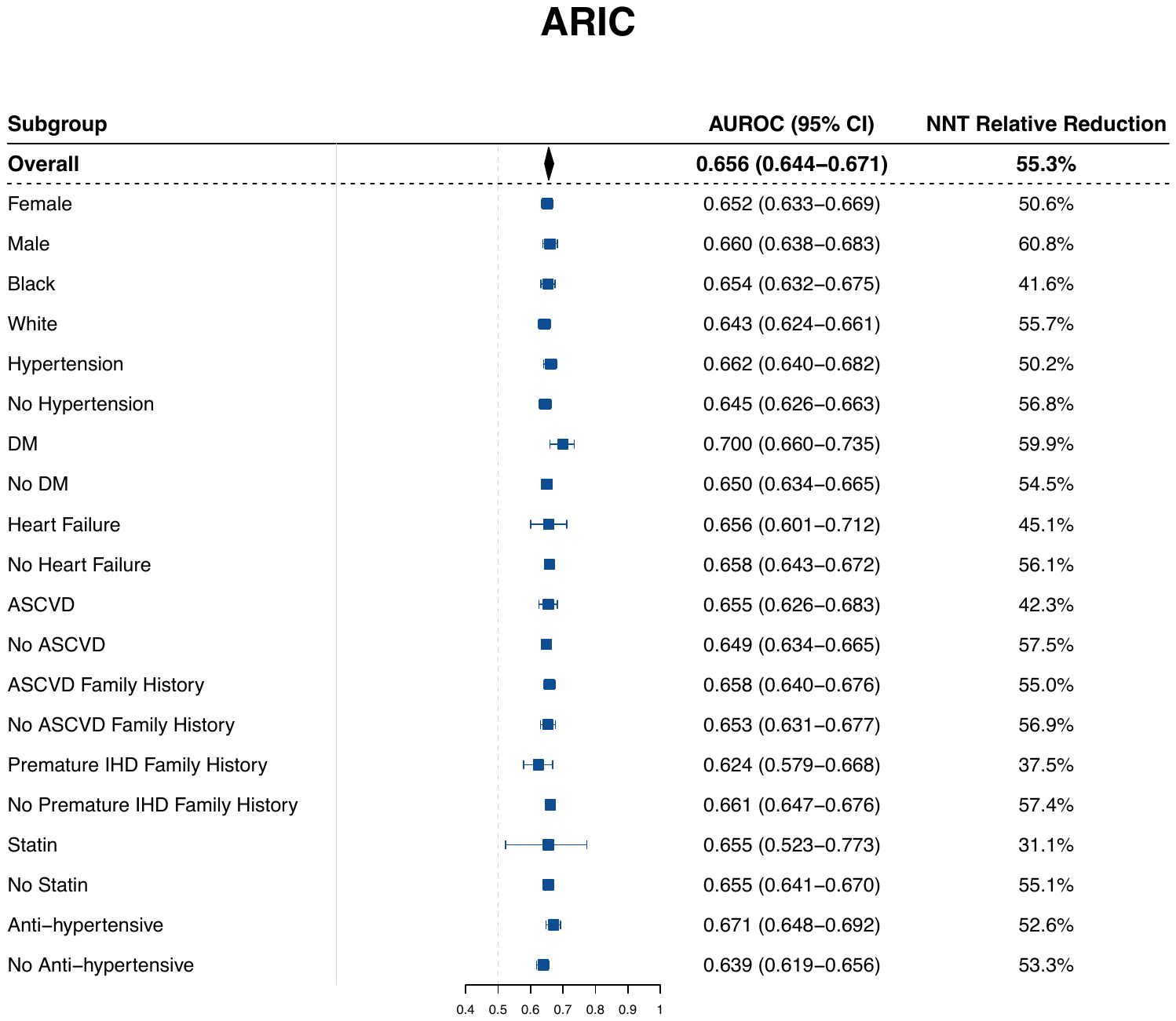

Abbreviations: ARIC, Atherosclerosis Risk in Communities; ASCVD, atherosclerotic cardiovascular disease; AUROC, area under the receiver operating characteristic curve; CI, confidence interval; DM, diabetes mellitus; IHD, ischemic heart disease; NNT, number needed to test.

### **eFigure 4. ARISE’s Performance and Number Needed to Test Relative Reduction Across Demographic and Clinical Subgroups in the CARDIA Cohort**

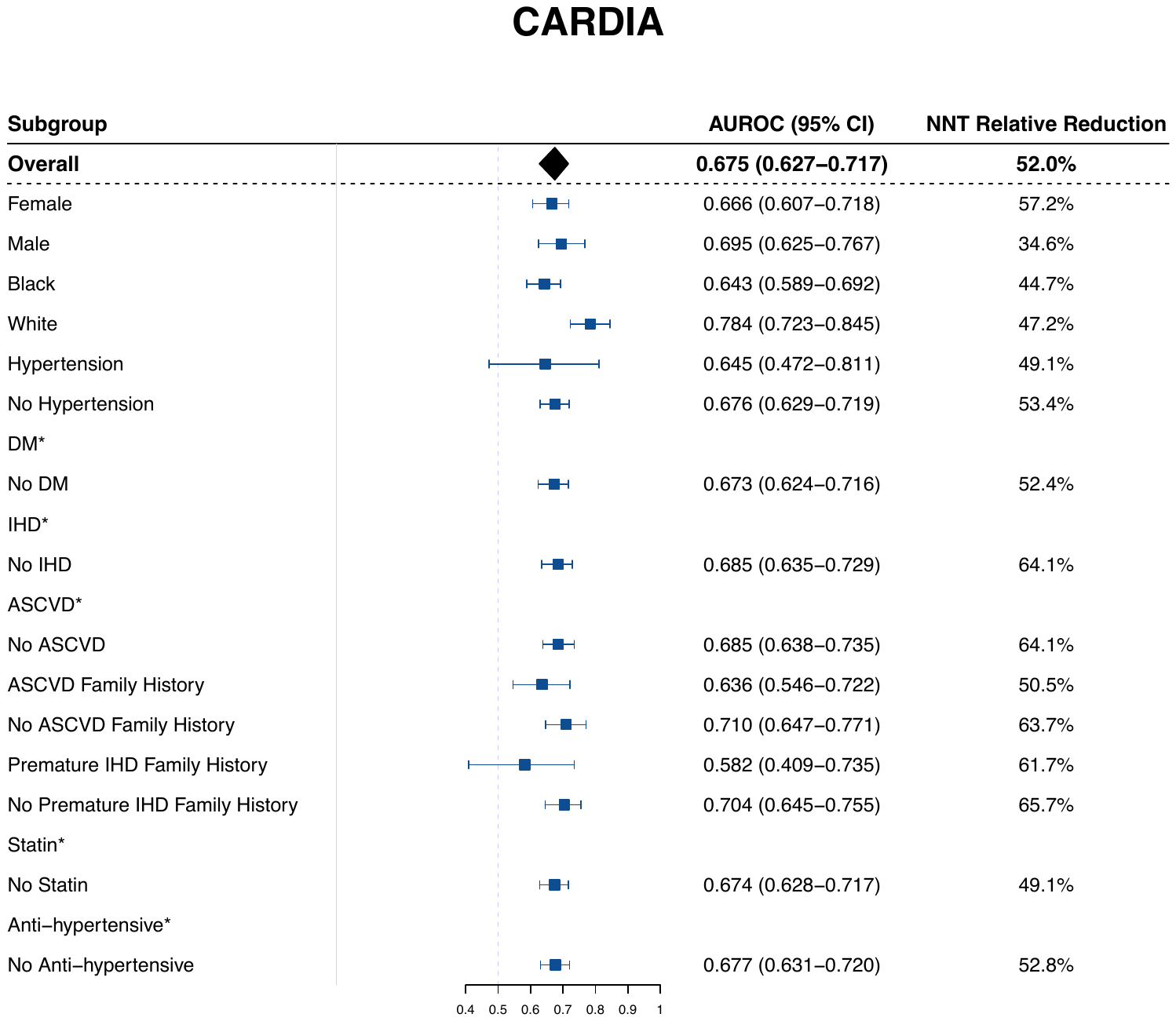

*Comprises of less than 10 participants with elevated Lp(a).

Abbreviations: ASCVD, atherosclerotic cardiovascular disease; AUROC, area under the receiver operating characteristic curve; CARDIA, Coronary Artery Risk Development in Young Adults; CI, confidence interval; DM, diabetes mellitus; IHD, ischemic heart disease; NNT, number needed to test.

### **eFigure 5. ARISE’s Performance and Number Needed to Test Relative Reduction Across Demographic and Clinical Subgroups in the MESA Cohort**

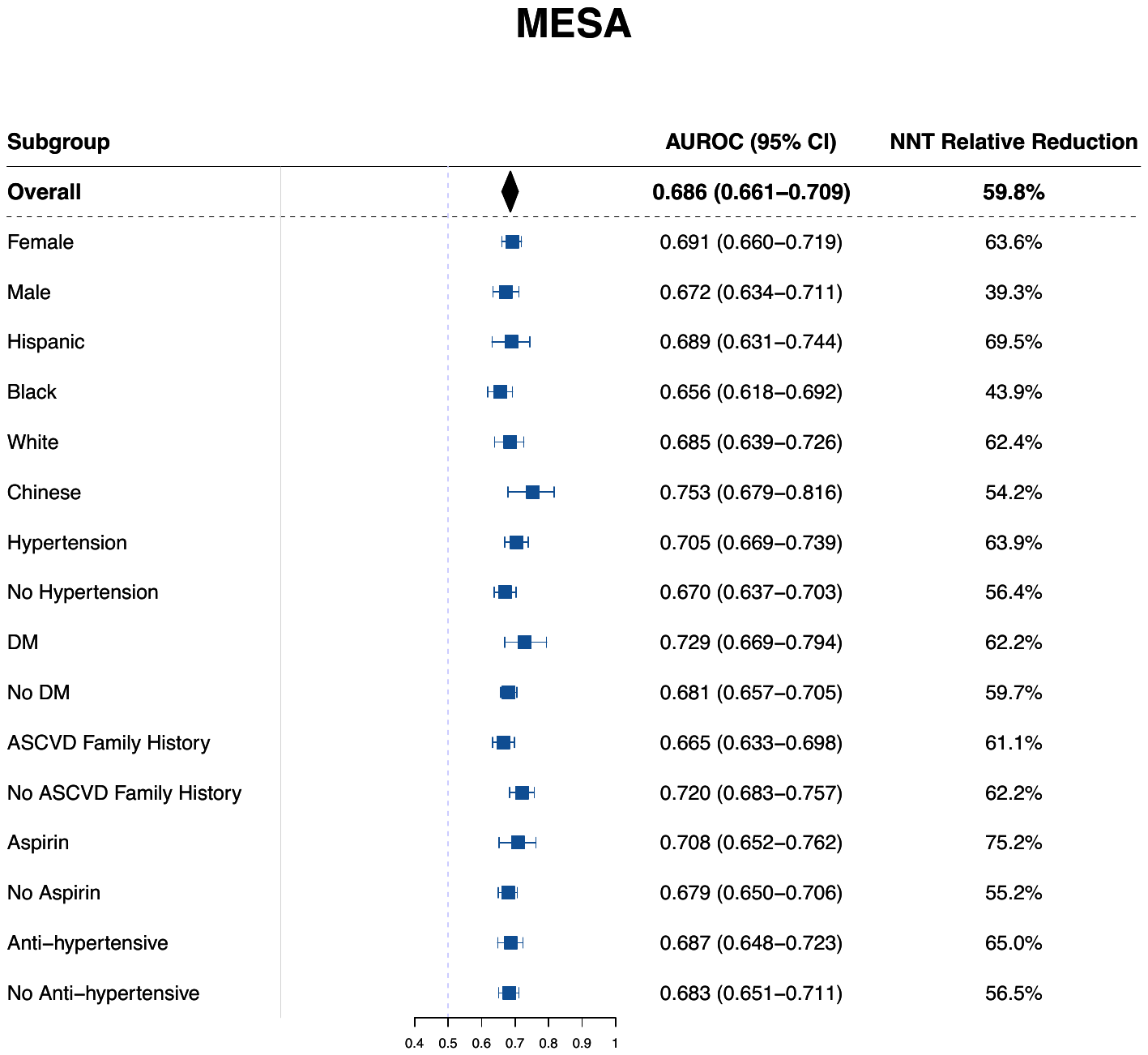

Abbreviations: ASCVD, atherosclerotic cardiovascular disease; AUROC, area under the receiver operating characteristic curve; CI, confidence interval; DM, diabetes mellitus; MESA, Multi-Ethnic Study of Atherosclerosis; NNT, number needed to test.

### **eTable 1.** **Definition of the Variables in the UK Biobank**

| **Coded Variable** | **Instances from UK Biobank** | |
| --- | --- | --- |
| **Medications** | | |
| **Medication Category** | **Medication Name** | |
| Aspirin | Aspirin 75mg, Nu-Seals 75mg, Aspirin, Isosorbide mononitrate+Aspirin, Dipyridamole+Aspirin, Caprin 75mg, Micropirin 75mg, Disprin CV 100mg, Enprin 75mg, Postmi 75mg, Angettes 75mg | |
| Statin | Simvastatin, Velastatin, Zocor, Simvador, Synvinolin, Fluvastatin, Lescol, Pravastatin, Eptastatin, Lipostat, Atorvastatin, Lipitor, Rosuvastatin, Crestor | |
| Lipid-lowering Therapies | Provided by the UK Biobank | |
| Anti-hypertensive Medications | Provided by the UK Biobank | |
| Insulin | Provided by the UK Biobank | |
| Hormone Replacement Therapy | Provided by the UK Biobank | |
| Contraceptive Pills | Provided by the UK Biobank | |
| **Inpatient Diagnoses** | | |
| **Condition** | **ICD-9 Code** | **ICD-10 Code** |
| Hypertension | 401, 4010, 4011, 4019, 402, 4020, 4021, 4029, 403, 4030, 4031, 4039, 404, 4040, 4041, 4049, 6420, 6422, 6427, 6429 | I10, I11, I110, I119, I12, I120, I129, I13, I130, I131, I132, I139, I674, O10, O100, O101, O102, O103, O109, O11 |
| Diabetes Mellitus | 250, 2500, 25000, 25001, 25009, 2501, 25010, 25011, 25019, 2502, 25020, 25021, 25029, 2503, 2504, 2505, 2506, 2507, 2509, 25090, 25091, 25099, 6480 | E10, E100, E101, E102, E103, E104, E105, E106, E107, E108, E109, E11, E110, E111, E112, E113, E114, E115, E116, E117, E118, E119, E12, E120, E121, E122, E123, E124, E125, E126, E127, E128, E129, E13, E130, E131, E132, E133, E134, E135, E136, E137, E138, E139, E14, E140, E141, E142, E143, E144, E145, E146, E147, E148, E149, O240, O241, O242, O243, O249 |
| Type 1 Diabetes Mellitus | 25001, 25011, 25021, 25091 | E10, E100, E101, E102, E103, E104, E105, E106, E107, E108, E109, O240 |
| Type 2 Diabetes Mellitus | 25000, 25010, 25020, 25090 | E11, E110, E111, E112, E113, E114, E115, E116, E117, E118, E119, O241 |
| Chronic Kidney Disease | 403, 4030, 4031, 4039, 404, 4040, 4041, 4049, 585, 5859, 6421, 6462 | I12, I120, I13, I130, I131, I132, I139, N18, N180, N181, N182, N183, N184, N185, N188, N189, Z49, Z490, Z491, Z492 |
| Ischemic Heart Disease | 410, 4109, 411, 4119, 412, 4129, 413, 4139, 414, 4140, 4148, 4149 | I20, I200, I208, I209, I21, I210, I211, I212, I213, I214, I219, I21X, I22, I220, I221, I228, I229, I23, I230, I231, I232, I233, I234, I235, I236, I238, I24, I240, I241, I248, I249, I25, I250, I251, I252, I255, I256, I258, I259, Z951, Z955 |
| Heart Failure | 428, 4280, 4281, 4289 | I110, I130, I132, I50, I500, I501, I509 |
| Ischemic Stroke | 433, 4330, 4331, 4332, 4333, 4338, 4339, 434, 4340, 4341, 4349, 435, 4359, 437, 4370, 4371 | G45, G450, G451, G452, G453, G454, G458, G459, I63, I630, I631, I632, I633, I634, I635, I638, I639, I64, I65, I650, I651, I652, I653, I658, I659, I66, I660, I661, I662, I663, I664, I668, I669, I672, I693, I694 |
| Peripheral Arterial Disease | 4402, 4442 | I702, I7020, I7021, I742, I743, I744 |
| **Inpatient Main/Primary Diagnoses** | | |
| **Hospitalization Main Diagnosis** | **ICD-9 Code** | **ICD-10 Code** |
| Ischemic Heart Disease | 410, 4109 | I200, I21, I210, I211, I212, I213, I214, I219, I21X, I22, I220, I221, I228, I229, I24, I248, I249, |
| Heart Failure | 428, 4280, 4281, 4289 | I110, I130, I132, I50, I500, I501, I509 |
| Ischemic Stroke | 433, 4330, 4331, 4332, 4333, 4338, 4339, 434, 4340, 4341, 4349, 435, 4359 | G45, G450, G451, G452, G453, G454, G458, G459, I63, I630, I631, I632, I633, I634, I635, I638, I639, I64 |
| Peripheral Arterial Disease | 4442 | I742, I743, I744 |
| **Inpatient Procedures** | | |
| **Procedure** | **OPCS-3 Code** | **OPCS-4 Code** |
| Coronary Artery Bypass Grafting | 3043 | K40, K401, K402, K403, K404, K408, K409, K41, K411, K412, K413, K414, K418, K419, K42, K421, K422, K423, K424, K428, K429, K43, K431, K432, K433, K434, K438, K439, K44, K441, K442, K448, K449, K45, K451, K452, K453, K454, K455, K456, K458, K459, K46, K461, K462, K463, K464, K465, K468, K469 |
| Percutaneous Coronary Intervention | None | K49, K491, K492, K493, K494, K498, K499, K50, K501, K502, K503, K504, K508, K509, K75, K751, K752, K753, K754, K758, K759 |
| Carotid Revascularization | None | L29, L291, L292, L293, L294, L295, L296, L297, L298, L299, L303, L31, L311, L313, L314, L318, L319 |
| Peripheral Arterial Disease Revascularization | 881, 8811 | L50, L501, L502, L503, L504, L505, L506, L508, L509, L51, L511, L512, L513, L514, L515, L516, L518, L519, L52, L521, L522, L528, L529, L532, L54, L541, L542, L544, L548, L549, L58, L581, L582, L583, L584, L585, L586, L587, L588, L589, L59, L591, L592, L593, L594, L595, L596, L597, L598, L599, L60, L601, L602, L603, L604, L608, L609, L622, L63, L631, L632, L633, L635, L638, L639, L66, L661, L662, L665, L667, L681, L682, L701, L71, L711, L712, L713, L714, L715, L716, L717, L718, L719 |

### **eTable 2. Baseline Characteristics of the UK Biobank Participants with and without Lp(a) Measurement**

| **Characteristic** | **Overall (N=502,468)** | **Lp(a) Measurement (N=456,815)** | **No Lp(a) Measurement (N=45,653)** |
| --- | --- | --- | --- |
| **Age (years)** | 57.2 (8.14) | 57.2 (8.14) | 57.0 (8.13) |
| **Female Sex** | 273359 (54.4%) | 247511 (54.2%) | 25848 (56.6%) |
| **Ethnicity** |  |  |  |
| White | 472668 (94.6%) | 430601 (94.7%) | 42067 (93.4%) |
| Black | 8060 (1.6%) | 7079 (1.6%) | 981 (2.2%) |
| South Asian | 9881 (2.0%) | 8846 (1.9%) | 1035 (2.3%) |
| Chinese | 1574 (0.3%) | 1418 (0.3%) | 156 (0.3%) |
| Mixed | 2956 (0.6%) | 2683 (0.6%) | 273 (0.6%) |
| Other | 4558 (0.9%) | 4052 (0.9%) | 506 (1.1%) |
| **Smoking** |  |  |  |
| Never | 273656 (54.8%) | 248944 (54.8%) | 24712 (54.9%) |
| Former | 173207 (34.7%) | 157990 (34.8%) | 15217 (33.8%) |
| Current | 52652 (10.5%) | 47559 (10.5%) | 5093 (11.3%) |
| **BMI (kg/m2)** | 27.4 (4.80) | 27.4 (4.78) | 27.6 (5.02) |
| **Vital Signs** |  |  |  |
| **Systolic Blood Pressure (mmHg)** | 138 (18.7) | 138 (18.6) | 138 (19.0) |
| **Diastolic Blood Pressure (mmHg)** | 82.2 (10.1) | 82.2 (10.1) | 82.2 (10.3) |
| **Heart Rate (beats per minute)** | 69.4 (11.3) | 69.3 (11.3) | 69.7 (11.4) |
| **Medical History** |  |  |  |
| **Hypertension** | 41223 (8.2%) | 37368 (8.2%) | 3855 (8.4%) |
| **DM** | 11470 (2.3%) | 10297 (2.3%) | 1173 (2.6%) |
| **T1DM** | 2297 (0.5%) | 2033 (0.4%) | 264 (0.6%) |
| **T2DM** | 9701 (1.9%) | 8722 (1.9%) | 979 (2.1%) |
| **CKD** | 1250 (0.2%) | 1122 (0.2%) | 128 (0.3%) |
| **IHD** | 21009 (4.2%) | 19036 (4.2%) | 1973 (4.3%) |
| **Heart Failure** | 2315 (0.5%) | 2069 (0.5%) | 246 (0.5%) |
| **Ischemic Stroke** | 3691 (0.7%) | 3329 (0.7%) | 362 (0.8%) |
| **PAD** | 651 (0.1%) | 583 (0.1%) | 68 (0.1%) |
| **CABG** | 3207 (0.6%) | 2912 (0.6%) | 295 (0.6%) |
| **PCI** | 5610 (1.1%) | 5089 (1.1%) | 521 (1.1%) |
| **Carotid Revascularization** | 310 (0.1%) | 278 (0.1%) | 32 (0.1%) |
| **PAD Revascularization** | 1239 (0.2%) | 1113 (0.2%) | 126 (0.3%) |
| **ASCVD** | 24893 (5.0%) | 22534 (4.9%) | 2359 (5.2%) |
| **Premature ASCVD** | 12702 (2.5%) | 11447 (2.5%) | 1255 (2.7%) |
| **Age at First ASCVD** | 57.0 (6.83) | 57.0 (6.81) | 56.7 (7.06) |
| **Family History of ASCVD** | 289356 (58.8%) | 263497 (58.9%) | 25859 (58.5%) |
| **Medication History** |  |  |  |
| **Aspirin** | 69635 (13.9%) | 63485 (13.9%) | 6150 (13.5%) |
| **Statin** | 83801 (16.7%) | 76292 (16.7%) | 7509 (16.4%) |
| **Lipid-lowering Therapy** | 88216 (17.8%) | 80723 (17.8%) | 7493 (18.1%) |
| **Anti-hypertensive Medication** | 105115 (21.3%) | 96182 (21.2%) | 8933 (21.6%) |
| **Insulin** | 5640 (1.1%) | 5059 (1.1%) | 581 (1.4%) |
| **Hormone Replacement Therapy** | 19619 (4.0%) | 17904 (4.0%) | 1715 (4.1%) |
| **Contraceptive Pill** | 6935 (1.4%) | 6310 (1.4%) | 625 (1.5%) |
| **Laboratory Values** |  |  |  |
| **Hemoglobin (g/dL)** | 14.2 (1.25) | 14.2 (1.25) | 14.2 (1.27) |
| **MCV (fL)** | 91.1 (4.60) | 91.1 (4.60) | 91.1 (4.65) |
| **WBC (10^9^/L)** | 6.89 (2.13) | 6.89 (2.13) | 6.92 (2.25) |
| **Platelet (10^9^/L)** | 252 (60.1) | 252 (60.1) | 252 (60.8) |
| **LDL-C (mmol/L)** | 3.56 (0.871) | 3.56 (0.871) | 3.59 (0.879) |
| **HDL-C (mmol/L)** | 1.45 (0.383) | 1.45 (0.383) | 1.47 (0.393) |
| **Total Cholesterol (mmol/L)** | 5.69 (1.15) | 5.69 (1.15) | 5.74 (1.15) |
| **Triglycerides (mmol/L)** | 1.75 (1.04) | 1.75 (1.04) | 1.73 (1.07) |
| **FBS (mmol/L)** | 5.13 (1.24) | 5.12 (1.24) | 5.17 (1.31) |
| **HbA1C (mmol/mol)** | 36.1 (6.80) | 36.1 (6.78) | 36.3 (7.01) |
| **ALT (U/L)** | 23.6 (14.5) | 23.6 (14.4) | 23.5 (16.4) |
| **AST (U/L)** | 26.3 (11.0) | 26.2 (10.8) | 26.4 (14.3) |
| **ALP (U/L)** | 83.8 (26.7) | 83.7 (26.6) | 84.1 (30.1) |
| **Direct Bilirubin (µmol/L)** | 1.71 (0.839) | 1.71 (0.839) | 1.70 (0.830) |
| **Total Bilirubin (µmol/L)** | 9.11 (4.43) | 9.11 (4.42) | 9.25 (4.55) |
| **C-Reactive Protein (mg/L)** | 2.65 (4.80) | 2.65 (4.79) | 2.69 (4.93) |
| **Calcium (mmol/L)** | 2.38 (0.0944) | 2.38 (0.0942) | 2.40 (0.101) |
| **Phosphate (mmol/L)** | 1.16 (0.162) | 1.16 (0.162) | 1.17 (0.163) |
| **Total Protein (g/L)** | 72.6 (4.13) | 72.5 (4.12) | 73.2 (4.51) |
| **Albumin (g/L)** | 45.2 (2.64) | 45.2 (2.63) | 45.4 (2.89) |
| **Vitamin D (nmol/L)** | 48.3 (21.3) | 48.3 (21.3) | 49.1 (21.2) |

Abbreviations: ASCVD, atherosclerotic cardiovascular disease; ALP, alkaline phosphatase; ALT, alanine aminotransferase; ASCVD, atherosclerotic cardiovascular disease; AST, aspartate aminotransferase; BMI, body mass index; CABG, coronary artery bypass grafting surgery; CKD, chronic kidney disease; DM, diabetes mellitus; FBS, fasting blood sugar; HbA1C, hemoglobin A1C; HDL-C, high-density lipoprotein cholesterol; IHD, ischemic heart disease; LDL-C, low-density lipoprotein cholesterol; MCV, mean corpuscular volume; PAD, peripheral arterial disease; PCI, percutaneous coronary intervention; T1DM, type 1 diabetes mellitus; T2DM, type 2 diabetes mellitus; WBC, white blood cells.

### **eTable 3. Baseline Characteristics of the UK Biobank Participants with Normal and High Lp(a)**

| **Characteristic** | **Overall (N=456,815)** | **Normal Lp(a) (N=399,463)** | **High Lp(a) (N=57,352)** |
| --- | --- | --- | --- |
| **Age (years)** | 57.2 (8.14) | 57.1 (8.18) | 58.0 (7.82) |
| **Female Sex** | 247511 (54.2%) | 213471 (53.4%) | 34040 (59.4%) |
| **Ethnicity** |  |  |  |
| White | 430601 (94.7%) | 376576 (94.7%) | 54025 (94.6%) |
| Black | 7079 (1.6%) | 5566 (1.4%) | 1513 (2.7%) |
| South Asian | 8846 (1.9%) | 8095 (2.0%) | 751 (1.3%) |
| Chinese | 1418 (0.3%) | 1346 (0.3%) | 72 (0.1%) |
| Mixed | 2683 (0.6%) | 2351 (0.6%) | 332 (0.6%) |
| Other | 4052 (0.9%) | 3650 (0.9%) | 402 (0.7%) |
| **Smoking** |  |  |  |
| Never | 248944 (54.8%) | 218127 (54.9%) | 30817 (54.0%) |
| Former | 157990 (34.8%) | 137753 (34.7%) | 20237 (35.5%) |
| Current | 47559 (10.5%) | 41565 (10.5%) | 5994 (10.5%) |
| **BMI (kg/m2)** | 27.4 (4.78) | 27.4 (4.78) | 27.5 (4.77) |
| **Vital Signs** |  |  |  |
| **Systolic Blood Pressure (mmHg)** | 138 (18.6) | 138 (18.6) | 139 (18.7) |
| **Diastolic Blood Pressure (mmHg)** | 82.2 (10.1) | 82.2 (10.1) | 82.4 (10.1) |
| **Heart Rate (beats per minute)** | 69.3 (11.3) | 69.4 (11.3) | 69.2 (11.3) |
| **Medical History** |  |  |  |
| **Hypertension** | 37368 (8.2%) | 31484 (7.9%) | 5884 (10.3%) |
| **DM** | 10297 (2.3%) | 8916 (2.2%) | 1381 (2.4%) |
| **T1DM** | 2033 (0.4%) | 1732 (0.4%) | 301 (0.5%) |
| **T2DM** | 8722 (1.9%) | 7559 (1.9%) | 1163 (2.0%) |
| **CKD** | 1122 (0.2%) | 933 (0.2%) | 189 (0.3%) |
| **IHD** | 19036 (4.2%) | 15265 (3.8%) | 3771 (6.6%) |
| **Acute IHD Hospitalization** |  |  |  |
| None | 449618 (98.4%) | 393790 (98.6%) | 55828 (97.3%) |
| 1 | 6551 (1.4%) | 5180 (1.3%) | 1371 (2.4%) |
| 2 | 589 (0.1%) | 446 (0.1%) | 143 (0.2%) |
| ≥3 | 57 (0.01%) | 47 (0.01%) | 10 (0.02%) |
| **Heart Failure** | 2069 (0.5%) | 1704 (0.4%) | 365 (0.6%) |
| **Acute Heart Failure Hospitalization** |  |  |  |
| None | 456239 (99.9%) | 398982 (99.9%) | 57257 (99.8%) |
| 1 | 540 (0.1%) | 449 (0.1%) | 91 (0.2%) |
| 2 | 35 (0.01%) | 31 (0.01%) | 4 (0.01%) |
| ≥3 | 1 (0.0%) | 1 (0.0%) | 0 (0%) |
| **Ischemic Stroke** | 3329 (0.7%) | 2752 (0.7%) | 577 (1.0%) |
| **Acute Ischemic Stroke Hospitalization** |  |  |  |
| None | 454231 (99.4%) | 397333 (99.5%) | 56898 (99.2%) |
| 1 | 2465 (0.5%) | 2027 (0.5%) | 438 (0.8%) |
| 2 | 116 (0.03%) | 100 (0.03%) | 16 (0.03%) |
| ≥3 | 3 (0.0%) | 3 (0.0%) | 0 (0%) |
| **PAD** | 583 (0.1%) | 454 (0.1%) | 129 (0.2%) |
| **Acute PAD Hospitalization** |  |  |  |
| None | 456583 (99.9%) | 399286 (100.0%) | 57297 (99.9%) |
| 1 | 231 (0.1%) | 176 (0.04%) | 55 (0.1%) |
| 2 | 1 (0.0%) | 1 (0.0%) | 0 (0%) |
| **CABG** | 2912 (0.6%) | 2204 (0.6%) | 708 (1.2%) |
| **CABG Procedures** |  |  |  |
| None | 453903 (99.4%) | 397259 (99.4%) | 56644 (98.8%) |
| 1 | 2896 (0.6%) | 2193 (0.5%) | 703 (1.2%) |
| 2 | 16 (0.0%) | 11 (0.0%) | 5 (0.0%) |
| **PCI** | 5089 (1.1%) | 3932 (1.0%) | 1157 (2.0%) |
| **PCI Procedures** |  |  |  |
| None | 451726 (98.9%) | 395531 (99.0%) | 56195 (98.0%) |
| 1 | 4653 (1.0%) | 3603 (0.9%) | 1050 (1.8%) |
| 2 | 408 (0.1%) | 311 (0.1%) | 97 (0.2%) |
| ≥3 | 28 (0.01%) | 18 (0.0%) | 10 (0.02%) |
| **Carotid Revascularization** | 278 (0.1%) | 211 (0.1%) | 67 (0.1%) |
| **Carotid Revascularization Procedures** |  |  |  |
| None | 456537 (99.9%) | 399252 (99.9%) | 57285 (99.9%) |
| 1 | 263 (0.1%) | 198 (0.0%) | 65 (0.1%) |
| 2 | 15 (0.0%) | 13 (0.0%) | 2 (0.0%) |
| **PAD Revascularization** | 1113 (0.2%) | 886 (0.2%) | 227 (0.4%) |
| **PAD Revascularization Procedures** |  |  |  |
| None | 455702 (99.8%) | 398577 (99.8%) | 57125 (99.6%) |
| 1 | 964 (0.2%) | 765 (0.2%) | 199 (0.3%) |
| 2 | 109 (0.02%) | 91 (0.02%) | 18 (0.03%) |
| ≥3 | 40 (0.01%) | 30 (0.01%) | 10 (0.02%) |
| **ASCVD** | 22534 (4.9%) | 18152 (4.5%) | 4382 (7.6%) |
| **10-year ASCVD Risk* (PCE, %)** | 8.40 (7.42) | 8.37 (7.44) | 8.63 (7.28) |
| **Premature ASCVD** | 11447 (2.5%) | 9131 (2.3%) | 2316 (4.0%) |
| **Age at First ASCVD** | 57.0 (6.81) | 57.0 (6.84) | 56.9 (6.65) |
| **Family History of ASCVD** | 263497 (58.9%) | 227403 (58.1%) | 36094 (64.2%) |
| **Medication History** |  |  |  |
| **Aspirin** | 63485 (13.9%) | 53703 (13.4%) | 9782 (17.1%) |
| **Statin** | 76292 (16.7%) | 62221 (15.6%) | 14071 (24.5%) |
| **Lipid-lowering Therapy** | 80723 (17.8%) | 65988 (16.7%) | 14735 (25.9%) |
| **Anti-hypertensive Medication** | 96182 (21.2%) | 81942 (20.7%) | 14240 (25.0%) |
| **Insulin** | 5059 (1.1%) | 4374 (1.1%) | 685 (1.2%) |
| **Hormone Replacement Therapy** | 17904 (4.0%) | 15877 (4.0%) | 2027 (3.6%) |
| **Contraceptive Pill** | 6310 (1.4%) | 5801 (1.5%) | 509 (0.9%) |
| **Laboratory Values** |  |  |  |
| **Hemoglobin (g/dL)** | 14.2 (1.25) | 14.2 (1.25) | 14.1 (1.20) |
| **MCV (fL)** | 91.1 (4.60) | 91.1 (4.61) | 91.1 (4.52) |
| **WBC (10^9^/L)** | 6.89 (2.13) | 6.88 (2.13) | 6.92 (2.06) |
| **Platelet (10^9^/L)** | 252 (60.1) | 252 (60.1) | 256 (60.0) |
| **LDL-C (mmol/L)** | 3.56 (0.871) | 3.53 (0.861) | 3.76 (0.915) |
| **HDL-C (mmol/L)** | 1.45 (0.383) | 1.44 (0.383) | 1.49 (0.380) |
| **Total Cholesterol (mmol/L)** | 5.69 (1.15) | 5.65 (1.13) | 5.96 (1.21) |
| **Triglycerides (mmol/L)** | 1.75 (1.04) | 1.76 (1.05) | 1.68 (0.951) |
| **FBS (mmol/L)** | 5.12 (1.24) | 5.12 (1.25) | 5.13 (1.20) |
| **HbA1C (mmol/mol)** | 36.1 (6.78) | 36.1 (6.78) | 36.5 (6.74) |
| **ALT (U/L)** | 23.6 (14.4) | 23.7 (14.7) | 22.7 (12.5) |
| **AST (U/L)** | 26.2 (10.8) | 26.3 (11.1) | 25.8 (8.95) |
| **ALP (U/L)** | 83.7 (26.6) | 83.4 (26.6) | 86.5 (26.5) |
| **Direct Bilirubin (µmol/L)** | 1.71 (0.839) | 1.72 (0.853) | 1.63 (0.735) |
| **Total Bilirubin (µmol/L)** | 9.11 (4.42) | 9.15 (4.45) | 8.86 (4.21) |
| **C-Reactive Protein (mg/L)** | 2.65 (4.79) | 2.63 (4.77) | 2.81 (4.93) |
| **Calcium (mmol/L)** | 2.38 (0.0942) | 2.38 (0.0939) | 2.39 (0.0955) |
| **Phosphate (mmol/L)** | 1.16 (0.162) | 1.16 (0.162) | 1.17 (0.159) |
| **Total Protein (g/L)** | 72.5 (4.12) | 72.5 (4.11) | 72.8 (4.14) |
| **Albumin (g/L)** | 45.2 (2.63) | 45.2 (2.62) | 45.1 (2.66) |
| **Vitamin D (nmol/L)** | 48.3 (21.3) | 48.3 (21.3) | 48.2 (21.3) |

*PCE was reported among participants without a history of ASCVD.

Abbreviations: ASCVD, atherosclerotic cardiovascular disease; ALP, alkaline phosphatase; ALT, alanine aminotransferase; ASCVD, atherosclerotic cardiovascular disease; AST, aspartate aminotransferase; BMI, body mass index; CABG, coronary artery bypass grafting surgery; CKD, chronic kidney disease; DM, diabetes mellitus; FBS, fasting blood sugar; HbA1C, hemoglobin A1C; HDL-C, high-density lipoprotein cholesterol; IHD, ischemic heart disease; LDL-C, low-density lipoprotein cholesterol; MCV, mean corpuscular volume; PAD, peripheral arterial disease; PCE, pooled cohort equation; PCI, percutaneous coronary intervention; T1DM, type 1 diabetes mellitus; T2DM, type 2 diabetes mellitus; WBC, white blood cells.

### **eTable 4. Performance of Different Machine Learning Models Using Available Features for Predicting Elevated Lp(a)**

| **Features** | **Number of Features** | **Cross-validation AUROC** | | |
| --- | --- | --- | --- | --- |
|  |  | **Logistic Regression** | **Extreme Gradient Boosting** | **TabNet** |
| **Demographics, Medications, Diagnosis Codes, Procedural Codes, Vital Signs, and Lab Measurements** | 4575 | 0.614 | 0.623 | 0.555 |
| **eTable 2 Features** | 68 | 0.667 | 0.670 | 0.666 |
| **ARISE’s Features** | 6 | 0.652 | 0.656 | 0.656 |

Abbreviations: ARISE, Algorithmic Risk Inspection for Screening Elevated Lp(a); AUROC, area under the receiver operating characteristic curve

### **eTable 5. Baseline Characteristics of Participants across UK Biobank, ARIC, CARDIA, and MESA Cohorts**

| **Characteristic** | **UK Biobank (N=456,815)** | **ARIC (N=14,484)** | **CARDIA (N=4,124)** | **MESA (N=4,672)** |
| --- | --- | --- | --- | --- |
| **Age (years)** | 57.2 (8.14) | 54.2 (5.77) | 30.0 (3.59) | 61.9 (10.4) |
| **Female Sex** | 247511 (54.2%) | 7867 (54.3%) | 2252 (54.6%) | 2453 (52.5%) |
| **Ethnicity** |  |  |  |  |
| White | 430601 (94.7%) | 10865 (75.0%) | 2135 (51.8%) | 1707 (36.5%) |
| Black | 7079 (1.6%) | 3619 (25.0%) | 1989 (48.2%) | 1344 (28.8%) |
| Chinese | 1418 (0.3%) | 0 (0%) | 0 (0%) | 559 (12.0%) |
| Other | 15581 (3.4%) | 0 (0%) | 0 (0%) | 0 (0%) |
| Hispanic | 0 (0%) | 0 (0%) | 0 (0%) | 1062 (22.7%) |
| **BMI (kg/m2)** | 27.4 (4.78) | 27.7 (5.33) | 26.0 (5.55) | 28.2 (5.44) |
| **DM** | 10297 (2.3%) | 1712 (11.8%) | 78 (1.9%) | 508 (10.9%) |
| **Hypertension** | 37368 (8.2%) | 4993 (34.6%) | 346 (8.5%) | 1950 (41.7%) |
| **ASCVD** | 22534 (4.9%) | 2314 (16.0%) | 335 (8.2%) | 0 (0%) |
| **Family History of ASCVD** | 263497 (58.9%) | 8212 (57.5%) | 993 (27.7%) | 2556 (58.3%) |
| **Statin** | 76292 (16.7%) | 84 (0.6%) | 11 (0.3%) | 0 (0%) |
| **Anti-hypertensive Medication** | 96182 (21.2%) | 4422 (30.7%) | 64 (1.6%) | 1548 (33.1%) |
| **LDL-C (mmol/L)** | 3.56 (0.871) | 3.57 (1.02) | 2.81 (0.824) | 3.09 (0.811) |
| **HDL-C (mmol/L)** | 1.45 (0.383) | 1.33 (0.440) | 1.38 (0.366) | 1.32 (0.389) |
| **Total Cholesterol (mmol/L)** | 5.69 (1.15) | 5.56 (1.09) | 4.61 (0.883) | 5.07 (0.917) |
| **Triglycerides (mmol/L)** | 1.75 (1.04) | 1.49 (1.02) | 0.910 (0.812) | 1.45 (0.956) |

Abbreviations: ARIC, Atherosclerosis Risk in Communities; ASCVD, atherosclerotic cardiovascular disease; BMI, body mass index; CARDIA, Coronary Artery Risk Development in Young Adults; HDL-C, high-density lipoprotein cholesterol; DM, diabetes mellitus; LDL-C, low-density lipoprotein cholesterol; MESA, Multi-Ethnic Study of Atherosclerosis.

### **eTable 6. Baseline Characteristics of ARIC Participants with Normal and High Lp(a)**

| **Characteristic** | **Overall (N=14,484)** | **Normal Lp(a) (N=12,823)** | **High Lp(a)** **(N=1,661)** | **P-value** |
| --- | --- | --- | --- | --- |
| **Age (years)** | 54.2 (5.77) | 54.2 (5.77) | 54.5 (5.72) | 0.046 |
| **Female Sex** | 7867 (54.3%) | 6811 (53.1%) | 1056 (63.6%) | <0.001 |
| **Ethnicity** |  |  |  | <0.001 |
| White | 10865 (75.0%) | 10000 (78.0%) | 865 (52.1%) |  |
| Black | 3619 (25.0%) | 2823 (22.0%) | 796 (47.9%) |  |
| **BMI (kg/m2)** | 27.7 (5.33) | 27.6 (5.27) | 28.2 (5.79) | <0.001 |
| **Hypertension** | 4993 (34.6%) | 4251 (33.3%) | 742 (44.8%) | <0.001 |
| **DM** | 1712 (11.8%) | 1488 (11.6%) | 224 (13.5%) | 0.027 |
| **IHD** | 613 (4.2%) | 512 (4.0%) | 101 (6.1%) | <0.001 |
| **Heart Failure** | 686 (4.8%) | 585 (4.6%) | 101 (6.2%) | 0.008 |
| **Stroke** | 670 (6.0%) | 577 (5.9%) | 93 (7.1%) | 0.087 |
| **PAD** | 588 (4.2%) | 480 (3.9%) | 108 (6.7%) | <0.001 |
| **CABG** | 224 (1.6%) | 186 (1.5%) | 38 (2.3%) | 0.014 |
| **PCI** | 356 (2.5%) | 299 (2.4%) | 57 (3.4%) | 0.009 |
| **Carotid Revascularization** | 39 (0.3%) | 36 (0.3%) | 3 (0.2%) | 0.618 |
| **PAD Revascularization** | 91 (0.6%) | 75 (0.6%) | 16 (1.0%) | 0.099 |
| **ASCVD** | 2314 (16.0%) | 1960 (15.3%) | 354 (21.3%) | <0.001 |
| **Family History of ASCVD** | 8212 (57.5%) | 7218 (57.0%) | 994 (61.1%) | 0.002 |
| **Family History of Premature IHD** | 1246 (8.8%) | 1080 (8.6%) | 166 (10.3%) | 0.027 |
| **Statin** | 84 (0.6%) | 64 (0.5%) | 20 (1.2%) | <0.001 |
| **Lipid-lowering Therapy** | 421 (2.9%) | 346 (2.7%) | 75 (4.6%) | <0.001 |
| **Anti-hypertensive Medication** | 4422 (30.7%) | 3748 (29.4%) | 674 (40.7%) | <0.001 |
| **LDL-C (mmol/L)** | 3.57 (1.02) | 3.51 (0.997) | 3.97 (1.07) | <0.001 |
| **HDL-C (mmol/L)** | 1.33 (0.440) | 1.32 (0.439) | 1.39 (0.446) | <0.001 |
| **Total Cholesterol (mmol/L)** | 5.56 (1.09) | 5.51 (1.07) | 5.98 (1.14) | <0.001 |
| **Triglycerides (mmol/L)** | 1.49 (1.02) | 1.51 (1.04) | 1.37 (0.816) | <0.001 |

Abbreviations: ARIC, Atherosclerosis Risk in Communities; ASCVD, atherosclerotic cardiovascular disease; BMI, body mass index; CABG, coronary artery bypass grafting surgery; DM, diabetes mellitus; HDL-C, high-density lipoprotein cholesterol; IHD, ischemic heart disease; LDL-C, low-density lipoprotein cholesterol; PAD, peripheral arterial disease; PCI, percutaneous coronary intervention.

### **eTable 7. Baseline Characteristics of CARDIA Participants with Normal and High Lp(a)**

| **Characteristic** | **Overall (N=4124)** | **Normal Lp(a) (N=3991)** | **High Lp(a) (N=133)** | **P-value** |
| --- | --- | --- | --- | --- |
| **Age (years)** | 30.0 (3.59) | 30.0 (3.60) | 30.0 (3.42) | 0.985 |
| **Female Sex** | 2252 (54.6%) | 2167 (54.3%) | 85 (63.9%) | 0.036 |
| **Ethnicity** |  |  |  | <0.001 |
| White | 2135 (51.8%) | 2112 (52.9%) | 23 (17.3%) |  |
| Black | 1989 (48.2%) | 1879 (47.1%) | 110 (82.7%) |  |
| **BMI (kg/m2)** | 26.0 (5.55) | 26.0 (5.50) | 27.8 (6.60) | 0.003 |
| **Hypertension** | 346 (8.5%) | 335 (8.5%) | 11 (8.3%) | >0.9 |
| **DM** | 78 (1.9%) | 75 (1.9%) | 3 (2.3%) | >0.9 |
| **IHD** | 335 (8.2%) | 329 (8.3%) | 6 (4.6%) | 0.178 |
| **Family History of ASCVD** | 993 (27.7%) | 952 (27.4%) | 41 (37.6%) | 0.026 |
| **Family History of Premature IHD** | 493 (14.9%) | 475 (14.8%) | 18 (18.9%) | 0.333 |
| **Lipid-lowering Therapy** | 11 (0.3%) | 10 (0.3%) | 1 (0.8%) | 0.804 |
| **Anti-hypertensive Medication** | 64 (1.6%) | 63 (1.6%) | 1 (0.8%) | 0.687 |
| **LDL-C (mmol/L)** | 2.81 (0.824) | 2.79 (0.818) | 3.33 (0.815) | <0.001 |
| **HDL-C (mmol/L)** | 1.38 (0.366) | 1.38 (0.366) | 1.42 (0.375) | 0.222 |
| **Total Cholesterol (mmol/L)** | 4.61 (0.883) | 4.59 (0.878) | 5.19 (0.838) | <0.001 |
| **Triglycerides (mmol/L)** | 0.910 (0.812) | 0.909 (0.819) | 0.942 (0.565) | 0.526 |

Abbreviations: ASCVD, atherosclerotic cardiovascular disease; BMI, body mass index; CARDIA, Coronary Artery Risk Development in Young Adults; DM, diabetes mellitus; HDL-C, high-density lipoprotein cholesterol; IHD, ischemic heart disease; LDL-C, low-density lipoprotein cholesterol.

### **eTable 8. Baseline Characteristics of MESA Participants with Normal and High Lp(a)**

| **Characteristic** | **Overall (N=4672)** | **Normal Lp(a) (N=4177)** | **High Lp(a) (N=495)** | **P-value** |
| --- | --- | --- | --- | --- |
| **Age (years)** | 61.9 (10.4) | 61.9 (10.4) | 62.1 (10.1) | 0.649 |
| **Female Sex** | 2453 (52.5%) | 2160 (51.7%) | 293 (59.2%) | 0.002 |
| **Ethnicity** |  |  |  | <0.001 |
| Caucasian | 1707 (36.5%) | 1571 (37.6%) | 136 (27.5%) |  |
| Black | 1344 (28.8%) | 1098 (26.3%) | 246 (49.7%) |  |
| Hispanic | 1062 (22.7%) | 981 (23.5%) | 81 (16.4%) |  |
| Chinese | 559 (12.0%) | 527 (12.6%) | 32 (6.5%) |  |
| **BMI (kg/m2)** | 28.2 (5.44) | 28.1 (5.44) | 28.6 (5.43) | 0.052 |
| **Hypertension** | 1950 (41.7%) | 1721 (41.2%) | 229 (46.3%) | 0.035 |
| **DM** | 508 (10.9%) | 444 (10.6%) | 64 (12.9%) | 0.142 |
| **T1DM** | 5 (0.1%) | 4 (0.1%) | 1 (0.2%) | >0.9 |
| **T2DM** | 374 (8.0%) | 326 (7.8%) | 48 (9.7%) | 0.17 |
| **ASCVD** | 0 | 0 | 0 | - |
| **Family History of ASCVD** | 2556 (58.3%) | 2269 (57.9%) | 287 (62.0%) | 0.101 |
| **Aspirin** | 775 (17.3%) | 679 (17.0%) | 96 (20.0%) | 0.114 |
| **Statin** | 0 | 0 | 0 | - |
| **Lipid-lowering Therapy** | 0 | 0 | 0 | - |
| **Anti-hypertensive Medication** | 1548 (33.1%) | 1357 (32.5%) | 191 (38.6%) | 0.008 |
| **Insulin** | 61 (1.3%) | 55 (1.3%) | 6 (1.2%) | >0.9 |
| **LDL-C (mmol/L)** | 3.09 (0.811) | 3.04 (0.793) | 3.55 (0.821) | <0.001 |
| **HDL-C (mmol/L)** | 1.32 (0.389) | 1.32 (0.387) | 1.35 (0.406) | 0.067 |
| **Total Cholesterol (mmol/L)** | 5.07 (0.917) | 5.02 (0.897) | 5.53 (0.953) | <0.001 |
| **Triglycerides (mmol/L)** | 1.45 (0.956) | 1.46 (0.979) | 1.35 (0.732) | 0.003 |

Abbreviations: ASCVD, atherosclerotic cardiovascular disease; BMI, body mass index; DM, diabetes mellitus; HDL-C, high-density lipoprotein cholesterol; IHD, ischemic heart disease; LDL-C, low-density lipoprotein cholesterol; MESA, Multi-Ethnic Study of Atherosclerosis; T1DM, type 1 diabetes mellitus; T2DM, type 2 diabetes mellitus.

### **eTable 9. ARISE’s Performance Measures Across Subgroups in the UK Biobank Held-out Test Set**

| **Subgroup** | **AUROC (95% CI)** | **AUPRC (95% CI)** | **Prevalence (95% CI)** | **PPV (95% CI)** | **Relative Reduction of NNT** | **Sensitivity (95% CI)** | **Specificity (95% CI)** | **NPV (95% CI)** |
| --- | --- | --- | --- | --- | --- | --- | --- | --- |
| **Female** | 0.655 (0.648-0.661) | 0.220 (0.213-0.227) | 0.138 (0.135-0.141) | 0.254 (0.250-0.258) | 45.7% | 0.235 (0.232-0.239) | 0.889 (0.887-0.892) | 0.879 (0.876-0.882) |
| **Male** | 0.650 (0.642-0.658) | 0.186 (0.178-0.194) | 0.110 (0.107-0.113) | 0.226 (0.222-0.230) | 51.1% | 0.205 (0.201-0.209) | 0.913 (0.910-0.915) | 0.902 (0.900-0.905) |
| **Black** | 0.651 (0.617-0.685) | 0.339 (0.296-0.394) | 0.219 (0.197-0.241) | 0.422 (0.397-0.448) | 48.2% | 0.221 (0.199-0.242) | 0.915 (0.901-0.930) | 0.807 (0.787-0.828) |
| **White** | 0.655 (0.649-0.660) | 0.205 (0.199-0.211) | 0.125 (0.123-0.128) | 0.242 (0.239-0.244) | 48.0% | 0.224 (0.221-0.226) | 0.899 (0.897-0.901) | 0.890 (0.888-0.892) |
| **South Asian** | 0.700 (0.654-0.741) | 0.180 (0.143-0.234) | 0.086 (0.073-0.100) | 0.217 (0.197-0.236) | 60.1% | 0.225 (0.206-0.245) | 0.923 (0.910-0.935) | 0.926 (0.914-0.939) |
| **Chinese** | 0.802 (0.655-0.908) | 0.113 (0.051-0.241) | 0.036 (0.014-0.058) | 0.048 (0.022-0.073) | 23.9% | 0.100 (0.065-0.135) | 0.925 (0.894-0.956) | 0.965 (0.943-0.986) |
| **Hypertension** | 0.661 (0.645-0.677) | 0.247 (0.228-0.269) | 0.156 (0.147-0.164) | 0.262 (0.252-0.272) | 40.8% | 0.404 (0.393-0.416) | 0.791 (0.781-0.800) | 0.878 (0.871-0.886) |
| **No Hypertension** | 0.653 (0.648-0.659) | 0.200 (0.194-0.206) | 0.123 (0.121-0.125) | 0.239 (0.236-0.242) | 48.6% | 0.203 (0.200-0.205) | 0.910 (0.908-0.912) | 0.891 (0.889-0.893) |
| **DM** | 0.646 (0.612-0.681) | 0.207 (0.175-0.248) | 0.127 (0.113-0.142) | 0.234 (0.216-0.253) | 45.7% | 0.327 (0.307-0.347) | 0.844 (0.829-0.860) | 0.896 (0.883-0.909) |
| **No DM** | 0.656 (0.650-0.661) | 0.206 (0.200-0.212) | 0.125 (0.123-0.128) | 0.243 (0.240-0.246) | 48.4% | 0.221 (0.218-0.223) | 0.902 (0.900-0.903) | 0.890 (0.888-0.892) |
| **IHD** | 0.641 (0.618-0.663) | 0.278 (0.251-0.308) | 0.189 (0.177-0.202) | 0.259 (0.245-0.273) | 26.8% | 0.634 (0.619-0.650) | 0.575 (0.560-0.591) | 0.871 (0.860-0.881) |
| **No IHD** | 0.653 (0.648-0.658) | 0.199 (0.194-0.205) | 0.123 (0.121-0.125) | 0.240 (0.237-0.242) | 48.8% | 0.195 (0.193-0.198) | 0.913 (0.911-0.915) | 0.890 (0.888-0.892) |
| **Heart Failure** | 0.658 (0.593-0.721) | 0.319 (0.240-0.419) | 0.195 (0.157-0.233) | 0.277 (0.234-0.321) | 29.7% | 0.475 (0.427-0.523) | 0.700 (0.656-0.744) | 0.846 (0.811-0.881) |
| **No Heart Failure** | 0.655 (0.650-0.660) | 0.205 (0.199-0.211) | 0.125 (0.123-0.127) | 0.242 (0.240-0.245) | 48.4% | 0.221 (0.219-0.224) | 0.901 (0.899-0.903) | 0.890 (0.888-0.892) |
| **ASCVD** | 0.640 (0.621-0.659) | 0.271 (0.246-0.299) | 0.185 (0.173-0.196) | 0.251 (0.239-0.264) | 26.5% | 0.626 (0.612-0.640) | 0.577 (0.563-0.592) | 0.872 (0.862-0.882) |
| **No ASCVD** | 0.653 (0.648-0.658) | 0.199 (0.193-0.204) | 0.122 (0.120-0.125) | 0.241 (0.238-0.244) | 49.2% | 0.192 (0.189-0.194) | 0.916 (0.914-0.918) | 0.890 (0.888-0.892) |
| **Premature ASCVD** | 0.636 (0.606-0.665) | 0.272 (0.244-0.311) | 0.188 (0.172-0.204) | 0.250 (0.232-0.268) | 24.9% | 0.617 (0.597-0.637) | 0.572 (0.552-0.592) | 0.866 (0.852-0.880) |
| **No Premature ASCVD** | 0.654 (0.649-0.660) | 0.202 (0.197-0.208) | 0.124 (0.122-0.126) | 0.242 (0.239-0.245) | 48.8% | 0.208 (0.205-0.211) | 0.908 (0.906-0.910) | 0.890 (0.888-0.892) |
| **Aspirin** | 0.667 (0.654-0.681) | 0.248 (0.233-0.265) | 0.151 (0.145-0.157) | 0.261 (0.253-0.268) | 42.0% | 0.410 (0.401-0.418) | 0.793 (0.786-0.800) | 0.883 (0.877-0.889) |
| **No Aspirin** | 0.651 (0.645-0.657) | 0.195 (0.189-0.201) | 0.121 (0.119-0.124) | 0.236 (0.233-0.239) | 48.5% | 0.186 (0.184-0.189) | 0.917 (0.915-0.919) | 0.891 (0.889-0.893) |
| **Statin** | 0.638 (0.627-0.649) | 0.261 (0.250-0.275) | 0.183 (0.177-0.189) | 0.260 (0.253-0.267) | 29.5% | 0.531 (0.523-0.539) | 0.660 (0.653-0.668) | 0.863 (0.857-0.868) |
| **No Statin** | 0.644 (0.638-0.650) | 0.179 (0.174-0.185) | 0.114 (0.112-0.116) | 0.224 (0.221-0.227) | 49.0% | 0.125 (0.123-0.128) | 0.944 (0.942-0.946) | 0.893 (0.891-0.896) |
| **Anti-hypertensive** | 0.658 (0.647-0.669) | 0.239 (0.228-0.253) | 0.152 (0.147-0.157) | 0.263 (0.257-0.269) | 42.4% | 0.363 (0.356-0.369) | 0.819 (0.813-0.824) | 0.878 (0.873-0.882) |
| **No Anti-hypertensive** | 0.649 (0.644-0.655) | 0.192 (0.185-0.198) | 0.119 (0.116-0.121) | 0.230 (0.227-0.233) | 48.4% | 0.176 (0.173-0.178) | 0.921 (0.919-0.923) | 0.892 (0.890-0.895) |
| **ASCVD Family History** | 0.651 (0.644-0.658) | 0.218 (0.211-0.225) | 0.136 (0.133-0.138) | 0.253 (0.249-0.257) | 46.5% | 0.248 (0.245-0.252) | 0.885 (0.882-0.888) | 0.883 (0.880-0.885) |
| **No ASCVD Family History** | 0.658 (0.649-0.666) | 0.185 (0.177-0.194) | 0.111 (0.108-0.115) | 0.224 (0.220-0.228) | 50.2% | 0.181 (0.177-0.185) | 0.921 (0.919-0.924) | 0.900 (0.897-0.903) |

Abbreviations: ASCVD, atherosclerotic cardiovascular disease; AUROC, area under the receiver operating characteristics curve; AUPRC, area under the precision-recall curve; DM, diabetes mellitus; IHD, ischemic heart disease; NNT, number needed to test; NPV, negative predictive value; PPV, positive predictive value.

### **eTable 10. ARISE’s Performance Measures Across Subgroups in the ARIC Cohort**

| **Subgroup** | **AUROC (95% CI)** | **AUPRC (95% CI)** | **Prevalence (95% CI)** | **PPV (95% CI)** | **Relative Reduction of NNT** | **Sensitivity (95% CI)** | **Specificity (95% CI)** | **NPV (95% CI)** |
| --- | --- | --- | --- | --- | --- | --- | --- | --- |
| **Female** | 0.652 (0.633-0.669) | 0.220 (0.203-0.242) | 0.134 (0.127-0.142) | 0.271 (0.262-0.281) | 50.6% | 0.209 (0.200-0.218) | 0.913 (0.907-0.919) | 0.882 (0.874-0.889) |
| **Male** | 0.660 (0.638-0.683) | 0.173 (0.153-0.199) | 0.091 (0.084-0.098) | 0.233 (0.223-0.243) | 60.8% | 0.195 (0.185-0.205) | 0.935 (0.930-0.941) | 0.920 (0.914-0.927) |
| **Black** | 0.654 (0.632-0.675) | 0.333 (0.304-0.362) | 0.220 (0.206-0.233) | 0.376 (0.361-0.392) | 41.6% | 0.245 (0.231-0.259) | 0.886 (0.875-0.896) | 0.806 (0.793-0.819) |
| **White** | 0.643 (0.624-0.661) | 0.135 (0.121-0.152) | 0.080 (0.075-0.085) | 0.180 (0.172-0.187) | 55.7% | 0.166 (0.159-0.173) | 0.934 (0.930-0.939) | 0.928 (0.924-0.933) |
| **Hypertension** | 0.662 (0.640-0.682) | 0.253 (0.228-0.282) | 0.149 (0.139-0.158) | 0.298 (0.285-0.311) | 50.2% | 0.236 (0.224-0.248) | 0.903 (0.895-0.911) | 0.871 (0.862-0.881) |
| **No Hypertension** | 0.645 (0.626-0.663) | 0.164 (0.148-0.183) | 0.097 (0.091-0.103) | 0.224 (0.216-0.233) | 56.8% | 0.179 (0.171-0.186) | 0.934 (0.929-0.939) | 0.914 (0.908-0.919) |
| **DM** | 0.700 (0.660-0.735) | 0.249 (0.209-0.303) | 0.131 (0.115-0.147) | 0.327 (0.304-0.349) | 59.9% | 0.219 (0.199-0.238) | 0.932 (0.920-0.944) | 0.888 (0.873-0.903) |
| **No DM** | 0.650 (0.634-0.665) | 0.194 (0.179-0.213) | 0.112 (0.107-0.118) | 0.247 (0.240-0.255) | 54.5% | 0.202 (0.195-0.209) | 0.922 (0.918-0.927) | 0.901 (0.896-0.906) |
| **Heart Failure** | 0.656 (0.601-0.712) | 0.254 (0.195-0.340) | 0.147 (0.121-0.174) | 0.268 (0.235-0.301) | 45.1% | 0.218 (0.187-0.249) | 0.897 (0.875-0.920) | 0.869 (0.844-0.894) |
| **No Heart Failure** | 0.658 (0.643-0.672) | 0.199 (0.185-0.217) | 0.114 (0.108-0.119) | 0.258 (0.251-0.266) | 56.1% | 0.205 (0.198-0.212) | 0.925 (0.920-0.929) | 0.901 (0.896-0.906) |
| **ASCVD** | 0.655 (0.626-0.683) | 0.246 (0.215-0.288) | 0.153 (0.138-0.168) | 0.265 (0.247-0.283) | 42.3% | 0.331 (0.311-0.350) | 0.835 (0.820-0.850) | 0.873 (0.860-0.887) |
| **No ASCVD** | 0.649 (0.634-0.665) | 0.185 (0.170-0.203) | 0.107 (0.102-0.113) | 0.253 (0.245-0.260) | 57.5% | 0.170 (0.163-0.177) | 0.940 (0.935-0.944) | 0.904 (0.899-0.909) |
| **ASCVD Family History** | 0.658 (0.640-0.676) | 0.210 (0.192-0.232) | 0.121 (0.114-0.128) | 0.269 (0.259-0.278) | 55.0% | 0.208 (0.199-0.217) | 0.922 (0.916-0.928) | 0.894 (0.888-0.901) |
| **No ASCVD Family History** | 0.653 (0.631-0.677) | 0.187 (0.166-0.212) | 0.104 (0.097-0.112) | 0.242 (0.231-0.253) | 56.9% | 0.202 (0.192-0.212) | 0.926 (0.920-0.933) | 0.909 (0.902-0.916) |
| **Premature IHD Family History** | 0.624 (0.579-0.668) | 0.202 (0.164-0.254) | 0.133 (0.114-0.152) | 0.213 (0.190-0.236) | 37.5% | 0.157 (0.136-0.177) | 0.911 (0.895-0.927) | 0.875 (0.857-0.894) |
| **No Premature IHD Family History** | 0.661 (0.647-0.676) | 0.200 (0.184-0.217) | 0.112 (0.106-0.117) | 0.263 (0.255-0.270) | 57.4% | 0.211 (0.204-0.218) | 0.925 (0.921-0.930) | 0.903 (0.898-0.908) |
| **Statin** | 0.655 (0.523-0.773) | 0.328 (0.213-0.543) | 0.238 (0.147-0.329) | 0.345 (0.244-0.447) | 31.1% | 0.950 (0.903-0.997) | 0.438 (0.331-0.544) | 0.966 (0.926-1.005) |
| **No Statin** | 0.655 (0.641-0.670) | 0.197 (0.183-0.213) | 0.114 (0.109-0.119) | 0.253 (0.246-0.260) | 55.1% | 0.194 (0.187-0.200) | 0.927 (0.922-0.931) | 0.900 (0.895-0.904) |
| **Anti-hypertensive** | 0.671 (0.648-0.692) | 0.269 (0.242-0.300) | 0.152 (0.142-0.163) | 0.321 (0.308-0.335) | 52.6% | 0.266 (0.253-0.279) | 0.899 (0.890-0.908) | 0.872 (0.862-0.882) |
| **No Anti-hypertensive** | 0.639 (0.619-0.656) | 0.159 (0.145-0.177) | 0.098 (0.092-0.104) | 0.210 (0.202-0.218) | 53.3% | 0.162 (0.155-0.169) | 0.934 (0.929-0.939) | 0.911 (0.905-0.916) |

Abbreviations: ARIC, Atherosclerosis Risk in Communities; ASCVD, atherosclerotic cardiovascular disease; AUROC, area under the receiver operating characteristics curve; AUPRC, area under the precision-recall curve; DM, diabetes mellitus; IHD, ischemic heart disease; NNT, number needed to test; NPV, negative predictive value; PPV, positive predictive value.

### **eTable 11. ARISE’s Performance Measures Across Subgroups in the CARDIA Cohort**

| **Subgroup** | **AUROC (95% CI)** | **AUPRC (95% CI)** | **Prevalence (95% CI)** | **PPV (95% CI)** | **Relative Reduction of NNT** | **Sensitivity (95% CI)** | **Specificity (95% CI)** | **NPV (95% CI)** |
| --- | --- | --- | --- | --- | --- | --- | --- | --- |
| **Female** | 0.666 (0.607-0.718) | 0.077 (0.053-0.125) | 0.038 (0.030-0.046) | 0.088 (0.077-0.100) | 57.2% | 0.071 (0.060-0.081) | 0.971 (0.965-0.978) | 0.964 (0.956-0.972) |
| **Male** | 0.695 (0.625-0.767) | 0.053 (0.036-0.085) | 0.026 (0.018-0.033) | 0.039 (0.030-0.048) | 34.6% | 0.042 (0.033-0.051) | 0.973 (0.966-0.980) | 0.975 (0.968-0.982) |
| **Black** | 0.643 (0.589-0.692) | 0.099 (0.072-0.143) | 0.055 (0.045-0.065) | 0.100 (0.087-0.113) | 44.7% | 0.064 (0.053-0.074) | 0.966 (0.959-0.974) | 0.946 (0.936-0.956) |
| **White** | 0.784 (0.723-0.845) | 0.029 (0.016-0.054) | 0.011 (0.006-0.015) | 0.020 (0.014-0.026) | 47.2% | 0.043 (0.035-0.052) | 0.977 (0.971-0.984) | 0.989 (0.985-0.994) |
| **Hypertension** | 0.645 (0.472-0.811) | 0.143 (0.025-0.349) | 0.032 (0.013-0.050) | 0.062 (0.037-0.088) | 49.1% | 0.091 (0.061-0.121) | 0.955 (0.933-0.977) | 0.970 (0.952-0.988) |
| **No Hypertension** | 0.676 (0.629-0.719) | 0.061 (0.046-0.087) | 0.032 (0.027-0.038) | 0.069 (0.061-0.077) | 53.4% | 0.058 (0.050-0.065) | 0.974 (0.969-0.979) | 0.969 (0.963-0.974) |
| **DM*** |  |  |  |  |  |  |  |  |
| **No DM** | 0.673 (0.624-0.716) | 0.063 (0.049-0.091) | 0.032 (0.027-0.038) | 0.068 (0.060-0.076) | 52.4% | 0.062 (0.054-0.069) | 0.972 (0.967-0.977) | 0.969 (0.963-0.974) |
| **IHD*** |  |  |  |  |  |  |  |  |
| **No IHD** | 0.685 (0.635-0.729) | 0.070 (0.053-0.111) | 0.033 (0.027-0.039) | 0.092 (0.083-0.101) | 64.1% | 0.065 (0.057-0.072) | 0.978 (0.974-0.983) | 0.968 (0.963-0.974) |
| **ASCVD*** |  |  |  |  |  |  |  |  |
| **No ASCVD** | 0.685 (0.638-0.735) | 0.070 (0.052-0.105) | 0.033 (0.027-0.039) | 0.092 (0.083-0.101) | 64.1% | 0.065 (0.057-0.072) | 0.978 (0.974-0.983) | 0.968 (0.963-0.974) |
| **ASCVD Family History** | 0.636 (0.546-0.722) | 0.074 (0.045-0.144) | 0.041 (0.029-0.054) | 0.083 (0.066-0.101) | 50.5% | 0.073 (0.057-0.089) | 0.965 (0.954-0.977) | 0.960 (0.948-0.972) |
| **No ASCVD Family History** | 0.710 (0.647-0.771) | 0.067 (0.045-0.116) | 0.026 (0.020-0.032) | 0.072 (0.062-0.082) | 63.7% | 0.074 (0.063-0.084) | 0.975 (0.969-0.981) | 0.975 (0.969-0.981) |
| **Premature IHD Family History** | 0.582 (0.409-0.735) | 0.086 (0.030-0.230) | 0.037 (0.020-0.053) | 0.095 (0.069-0.121) | 61.7% | 0.111 (0.083-0.139) | 0.960 (0.943-0.977) | 0.966 (0.950-0.982) |
| **No Premature IHD Family History** | 0.704 (0.645-0.755) | 0.064 (0.045-0.105) | 0.027 (0.021-0.033) | 0.080 (0.070-0.090) | 65.7% | 0.078 (0.068-0.088) | 0.975 (0.969-0.981) | 0.974 (0.968-0.980) |
| **Statin*** |  |  |  |  |  |  |  |  |
| **No Statin** | 0.674 (0.628-0.717) | 0.061 (0.048-0.088) | 0.032 (0.027-0.038) | 0.063 (0.056-0.070) | 49.1% | 0.053 (0.046-0.060) | 0.974 (0.969-0.979) | 0.969 (0.963-0.974) |
| **Anti-hypertensive*** |  |  |  |  |  |  |  |  |
| **No Anti-hypertensive** | 0.677 (0.631-0.720) | 0.064 (0.048-0.092) | 0.033 (0.027-0.038) | 0.069 (0.061-0.077) | 52.8% | 0.061 (0.053-0.068) | 0.972 (0.967-0.978) | 0.969 (0.963-0.974) |

*Comprises of less than 10 participants with elevated Lp(a).

Abbreviations: ASCVD, atherosclerotic cardiovascular disease; AUROC, area under the receiver operating characteristics curve; AUPRC, area under the precision-recall curve; CARDIA, Coronary Artery Risk Development in Young Adults; DM, diabetes mellitus; IHD, ischemic heart disease; NNT, number needed to test; NPV, negative predictive value; PPV, positive predictive value.

### **eTable 12. ARISE’s Performance Measures Across Subgroups in the MESA Cohort**

| **Subgroup** | **AUROC (95% CI)** | **AUPRC (95% CI)** | **Prevalence (95% CI)** | **PPV (95% CI)** | **Relative Reduction of NNT** | **Sensitivity (95% CI)** | **Specificity (95% CI)** | **NPV (95% CI)** |
| --- | --- | --- | --- | --- | --- | --- | --- | --- |
| **Female** | 0.691 (0.660-0.719) | 0.232 (0.197-0.277) | 0.119 (0.107-0.132) | 0.329 (0.310-0.347) | 63.6% | 0.078 (0.068-0.089) | 0.978 (0.972-0.984) | 0.887 (0.874-0.899) |
| **Male** | 0.672 (0.634-0.711) | 0.162 (0.133-0.202) | 0.091 (0.079-0.103) | 0.150 (0.135-0.165) | 39.3% | 0.030 (0.023-0.037) | 0.983 (0.978-0.988) | 0.910 (0.898-0.922) |
| **Hispanic** | 0.689 (0.631-0.744) | 0.161 (0.111-0.242) | 0.076 (0.060-0.092) | 0.250 (0.224-0.276) | 69.5% | 0.062 (0.047-0.076) | 0.985 (0.977-0.992) | 0.927 (0.911-0.943) |
| **Black** | 0.656 (0.618-0.692) | 0.299 (0.255-0.358) | 0.183 (0.162-0.204) | 0.327 (0.301-0.352) | 43.9% | 0.065 (0.052-0.078) | 0.970 (0.961-0.979) | 0.822 (0.802-0.843) |
| **White** | 0.685 (0.639-0.726) | 0.152 (0.120-0.205) | 0.080 (0.067-0.093) | 0.212 (0.193-0.232) | 62.4% | 0.051 (0.041-0.062) | 0.983 (0.977-0.990) | 0.923 (0.910-0.936) |
| **Chinese** | 0.753 (0.679-0.816) | 0.125 (0.079-0.215) | 0.057 (0.038-0.077) | 0.125 (0.098-0.152) | 54.2% | 0.031 (0.017-0.046) | 0.987 (0.977-0.996) | 0.944 (0.925-0.963) |
| **Hypertension** | 0.705 (0.669-0.739) | 0.249 (0.207-0.298) | 0.117 (0.103-0.132) | 0.326 (0.305-0.346) | 63.9% | 0.061 (0.051-0.072) | 0.983 (0.977-0.989) | 0.887 (0.873-0.901) |
| **No Hypertension** | 0.670 (0.637-0.703) | 0.170 (0.141-0.209) | 0.098 (0.087-0.109) | 0.224 (0.208-0.240) | 56.4% | 0.056 (0.048-0.065) | 0.979 (0.973-0.984) | 0.905 (0.894-0.916) |
| **DM** | 0.729 (0.669-0.794) | 0.283 (0.200-0.400) | 0.126 (0.097-0.155) | 0.333 (0.292-0.374) | 62.2% | 0.047 (0.028-0.065) | 0.986 (0.976-0.997) | 0.878 (0.849-0.906) |
| **No DM** | 0.681 (0.657-0.705) | 0.193 (0.167-0.224) | 0.104 (0.094-0.113) | 0.257 (0.244-0.271) | 59.7% | 0.060 (0.053-0.068) | 0.980 (0.976-0.984) | 0.900 (0.891-0.909) |
| **ASCVD Family History** | 0.665 (0.633-0.698) | 0.205 (0.173-0.247) | 0.112 (0.100-0.125) | 0.288 (0.271-0.306) | 61.1% | 0.052 (0.044-0.061) | 0.984 (0.979-0.989) | 0.891 (0.879-0.903) |
| **No ASCVD Family History** | 0.720 (0.683-0.757) | 0.203 (0.162-0.257) | 0.096 (0.083-0.110) | 0.255 (0.235-0.275) | 62.2% | 0.068 (0.057-0.080) | 0.979 (0.972-0.985) | 0.908 (0.895-0.921) |
| **Aspirin** | 0.708 (0.652-0.762) | 0.268 (0.204-0.369) | 0.124 (0.101-0.147) | 0.500 (0.465-0.535) | 75.2% | 0.062 (0.045-0.080) | 0.991 (0.985-0.998) | 0.882 (0.859-0.905) |
| **No Aspirin** | 0.679 (0.650-0.706) | 0.191 (0.164-0.224) | 0.104 (0.094-0.114) | 0.232 (0.218-0.245) | 55.2% | 0.057 (0.050-0.065) | 0.978 (0.973-0.983) | 0.900 (0.890-0.909) |
| **Anti-hypertensive** | 0.687 (0.648-0.723) | 0.245 (0.201-0.304) | 0.123 (0.107-0.140) | 0.353 (0.329-0.377) | 65.0% | 0.063 (0.051-0.075) | 0.984 (0.977-0.990) | 0.882 (0.866-0.898) |
| **No Anti-hypertensive** | 0.683 (0.651-0.711) | 0.180 (0.154-0.217) | 0.097 (0.087-0.108) | 0.224 (0.209-0.238) | 56.5% | 0.056 (0.048-0.064) | 0.979 (0.974-0.984) | 0.906 (0.896-0.916) |

Abbreviations: ASCVD, atherosclerotic cardiovascular disease; AUROC, area under the receiver operating characteristics curve; AUPRC, area under the precision-recall curve; DM, diabetes mellitus; MESA, Multi-Ethnic Study of Atherosclerosis; NNT, number needed to test; NPV, negative predictive value; PPV, positive predictive value.

### **eTable 13. Baseline Characteristics of the UK Biobank Participants Included in the Exploratory Survival Analysis**

| **Characteristic** | **Overall (N=137,016)** | **Lp(a) Measurement (N=91,363)** | **No Lp(a) Measurement (N=45,653)** |
| --- | --- | --- | --- |
| **Age* (years)** | 65.1 (8.14) | 65.0 (8.14) | 65.2 (8.15) |
| **Female Sex** | 75354 (55.0%) | 49506 (54.2%) | 25848 (56.6%) |
| **Ethnicity** |  |  |  |
| White | 128266 (94.3%) | 86199 (94.8%) | 42067 (93.4%) |
| Black | 2388 (1.8%) | 1407 (1.5%) | 981 (2.2%) |
| South Asian | 2783 (2.0%) | 1748 (1.9%) | 1035 (2.3%) |
| Chinese | 432 (0.3%) | 276 (0.3%) | 156 (0.3%) |
| Mixed | 815 (0.6%) | 542 (0.6%) | 273 (0.6%) |
| Other | 1301 (1.0%) | 795 (0.9%) | 506 (1.1%) |
| **Smoking** |  |  |  |
| Never | 74534 (54.8%) | 49822 (54.8%) | 24712 (54.9%) |
| Former | 46655 (34.3%) | 31438 (34.6%) | 15217 (33.8%) |
| Current | 14727 (10.8%) | 9634 (10.6%) | 5093 (11.3%) |
| **BMI (kg/m^2^)** | 27.5 (4.87) | 27.4 (4.79) | 27.6 (5.02) |
| **Hypertension** | 11295 (8.2%) | 7440 (8.1%) | 3855 (8.4%) |
| **DM** | 3241 (2.4%) | 2068 (2.3%) | 1173 (2.6%) |
| **Heart Failure** | 656 (0.5%) | 410 (0.4%) | 246 (0.5%) |
| **ASCVD** | 6856 (5.0%) | 4497 (4.9%) | 2359 (5.2%) |
| **Premature ASCVD** | 3525 (2.6%) | 2270 (2.5%) | 1255 (2.7%) |
| **Family History of ASCVD** | 78601 (58.8%) | 52742 (58.9%) | 25859 (58.5%) |
| **Aspirin** | 18658 (13.6%) | 12508 (13.7%) | 6150 (13.5%) |
| **Statin** | 22580 (16.5%) | 15071 (16.5%) | 7509 (16.4%) |
| **Anti-hypertensive Medication** | 28195 (21.4%) | 19262 (21.3%) | 8933 (21.6%) |
| **Insulin** | 1599 (1.2%) | 1018 (1.1%) | 581 (1.4%) |
| **LDL-C (mmol/L)** | 3.56 (0.870) | 3.55 (0.869) | 3.59 (0.879) |
| **HDL-C (mmol/L)** | 1.45 (0.384) | 1.45 (0.382) | 1.47 (0.393) |
| **Total Cholesterol (mmol/L)** | 5.69 (1.14) | 5.69 (1.14) | 5.74 (1.15) |
| **Triglycerides (mmol/L)** | 1.75 (1.04) | 1.75 (1.04) | 1.73 (1.07) |
| **FBS (mmol/L)** | 5.13 (1.24) | 5.12 (1.23) | 5.17 (1.31) |
| **UK Biobank Study Visit^#^** |  |  |  |
| First | 133479 (97.4%) | 87826 (96.1%) | 45653 (100.0%) |
| Second | 3537 (2.6%) | 3537 (3.9%) | 0 (0%) |
| **ARISE Score** | 0.10 [0.09, 0.15] | 0.11 [0.08, 0.16] | 0.10 [0.10, 0.13] |
| **All-cause Mortality*** | 4406 (3.3%) | 2860 (3.2%) | 1546 (3.5%) |
| **Cardiovascular Mortality*** | 568 (0.4%) | 352 (0.4%) | 216 (0.5%) |
| **IHD*** | 1407 (1.0%) | 948 (1.0%) | 459 (1.0%) |
| **Stroke*** | 1198 (0.9%) | 817 (0.9%) | 381 (0.8%) |
| **MACE*** | 6661 (5.1%) | 4397 (5.0%) | 2264 (5.2%) |
| **Follow-up* (years)** | 4.2 [4.0, 4.5] | 4.2 [3.9, 4.7] | 4.2 [4.2, 4.2] |

*The variable is reported using Lp(a) assay date as the reference.

#Characteristics other than * variables were reported at the time of blood sample drawing for Lp(a) assay among those with Lp(a) measurement and at the time of the first UK Biobank study visit for those without Lp(a) measurement.

Abbreviations: ARISE, Algorithmic Risk Inspection for Screening Elevated Lp(a); ASCVD, atherosclerotic cardiovascular disease; BMI, body mass index; DM, diabetes mellitus; FBS, fasting blood sugar; HDL-C, high-density lipoprotein cholesterol; LDL-C, low-density lipoprotein cholesterol.
